# Optimizing functional connectivity scanning conditions for predicting autistic traits

**DOI:** 10.1101/2025.01.14.24319457

**Authors:** Corey Horien, Francesca Mandino, Abigail S. Greene, Xilin Shen, Kelly Powell, Angelina Vernetti, David O’Connor, Brendan D. Adkinson, Link Tejavibulya, James C. McPartland, Fred R. Volkmar, Marvin Chun, Katarzyna Chawarska, Evelyn M.R. Lake, Monica D. Rosenberg, Theodore Satterthwaite, Dustin Scheinost, Emily S. Finn, R. Todd Constable

## Abstract

Autism is a heterogeneous condition, and functional magnetic resonance imaging-based studies have advanced understanding of neurobiological correlates of autistic features. Little work has focused on the optimal brain states to reveal brain-phenotype relationships. Using connectome-based predictive modelling, we interrogated four datasets to determine scanning conditions that boost prediction of clinically relevant phenotypes and assess generalizability. In dataset one, a sample of youth with autism and neurotypical participants (n = 63), we found that a sustained attention task resulted in high prediction performance of autistic traits compared to a free-viewing social attention task and a resting-state condition. In dataset two (n = 25), we observed the predictive network model of autistic traits generated from the sustained attention task generalized to predict measures of attention in neurotypical adults. In datasets three and four, we determined the same predictive network model further generalized to predict measures of social responsiveness in the Autism Brain Imaging Data Exchange (n = 229) and the Healthy Brain Network (n = 643). Our data suggest an in-scanner sustained attention challenge can help delineate robust markers of autistic traits.

## Introduction

Autism spectrum disorder (referred to as “autism” hereafter) affects approximately 1% of children around the world^1^ and is characterized by difficulties with social communication and interaction, restricted and repetitive behaviors, and sensory atypicalities^2^. There is a need to better appreciate the neurobiological correlates of autistic traits in youth, which will help improve understanding of the condition and might aid potential clinical utility. Furthermore, there is a growing movement to characterize conditions like autism along dimensions of function^3–7^.

There are numerous approaches to characterize the brain-based correlates of autism traits using functional magnetic resonance imaging (fMRI) connectivity data, in which measures of similarity of the blood-oxygen-level-dependent (BOLD) signal are computed between different regions of interest^8^. In particular, prediction-based studies—using functional connectivity data to predict a phenotype—have proven promising. For instance, case-control studies have focused on classifying those with autism compared to neurotypical participants, showing that high prediction accuracy can be achieved on the basis of functional connectivity differences^9–18^. Another approach predicts continuous measures of a phenotype (a symptom scale or a behavioral test score)^18–21^. One method of dimensional prediction is connectome-based predictive modelling (CPM)^22,23^, which seeks to identify the functional connections most strongly predictive of a given phenotype. Groups using CPM in autism samples have identified brain correlates of clinician-rated autism symptoms^24,25^, and other traits, such as behavioral inhibition^26^, social responsiveness^24,27^, and attentional states^28^. Successful models tend to comprise distributed networks, spanning cortical, subcortical, and cerebellar regions. In particular, association cortices tend to be important in successful predictions, especially when the phenotypes relate to attention^24,28^. In addition, the most successful CPM models, while complex, consistently tend to comprise edges from about 5-10 percent of the connectome. This perhaps points to an optimal number of edges that might be involved in mediating brain-behavior relationships in those with autism.

Nevertheless, which conditions yield the best predictive modeling performance is still largely understudied. Most studies have typically focused on resting-state fMRI, in which participants rest quietly in the scanner. However, in neurotypical participants, the importance of scanning condition (e.g., ‘brain state’) is being recognized^29–32^ for prediction of various phenotypes, including intelligence^33–35^, attention^36,37^, working memory^38,39^, personality traits^40^, cognition and emotion scores^41^, as well as for emphasizing individual differences in connectivity patterns^42^. These studies suggest that predicting out-of-scanner phenotypes using connectivity measured during task performance tend to increase prediction accuracy, particularly when the task probes some aspect of the out-of-scanner item of interest (e.g., memory tasks in the scanner tend to result in higher prediction of memory performance outside the scanner^38^).

In addition, there are a number of elegant studies showing that in-scanner attention tasks can be used to inform the neurobiological organization of autism^43–47^. There are also other brain imaging studies suggesting an overlap between the functional networks mediating ADHD and autism^24,48^. At a behavioral level, the co-occurrence of autism and attention-deficit/hyperactivity disorder (ADHD) symptoms has long been acknowledged^49–52^.

Motivated by the importance of tasks in assessing phenotypes, as well as the importance of attention in autism, here we consider brain state-associated improvements in prediction performance in a sample of youth with autism and neurotypical participants. Using data from three different scanning conditions—a task requiring sustained attention, a task requiring selective social attention (SSA), and resting-state data—we applied CPM to probe brain-behavior relationships. Specifically, the gradual onset continuous performance task (gradCPT)^36,53,54^ tests the ability to sustain attention to constantly changing stimuli. The SSA task captures the ability to process dynamic, multimodal faces presented one-by-one within a complex visual scene. One of the best replicated eye-tracking biomarkers in autism^55–59^, the SSA is devoid of any narrative content or interpersonal interaction and was designed such that speech (SP) and eye contact (EC) were varied. The task design was constructed to determine the effect of each condition on prediction performance.

We hypothesized that consistent with the social features of autism, prediction performance of autistic traits would be highest in the SSA task and would increase with the presence of increased social cues. Specifically, we expected that the condition containing both eye contact and speech (EC+SP+) would yield the strongest prediction performance. We hypothesized that the next highest prediction performance would result from the sustained attention task, due to the restricted and repetitive behaviors observed in autism, and that both tasks would outperform resting-state data. To determine if results were robust, we used three other datasets to determine if successful models can generalize to external samples. One of the datasets was used to assess the model’s generalizability in predicting performance on an attention task; the other two datasets were used to assess prediction of other autistic features.

## Results

### Overview

Four samples were used in this work (Figure 1). The first dataset comprised 63 subjects from a sample described previously (mean age = 11.7 years, st. dev. = 2.8 years; 29 females; mean IQ = 107.8, st. dev. = 15.1; Supplemental Table 1)^28,60^. Twenty participants had autism; 11 other participants had a neurodevelopmental condition (five had attention-deficit/hyperactivity disorder (ADHD), two had anxiety disorder, and four were classified as belonging to the broader autism phenotype)^61^. Five of the participants with autism diagnoses had a co-occurring ADHD diagnosis; this was not significantly different compared to those without autism (*X*^2^ (1, *N* = 60) = 1.64 (Yates correction), *P* = 0.2001). Hereafter, we refer to this dataset as the “Yale youth sample”. It should be kept in mind that the sample comprised both a mixture of those with autism and those without, and there are only 20 participants with an autism diagnosis. Autism symptoms were scored using the Autism Diagnostic Observation Schedule-2 (ADOS-2)^62^.

**Figure 1.**
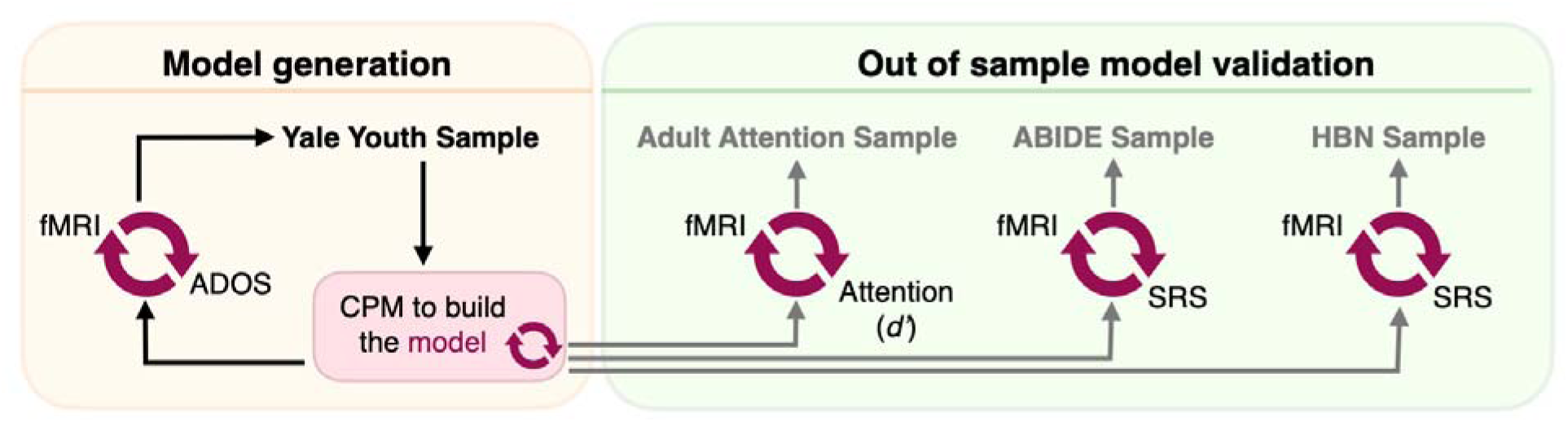
An overview of the datasets used in this study. The Yale youth sample was the first dataset used. FMRI connectivity data from different scanning conditions (task and rest) were used to generate connectome-based predictive models of Autism Diagnostic Observation Schedule (ADOS) scores (red circular arrows denote a brain-behavior predictive model). A summary predictive model was then generated and applied to the adult attention sample. The goal of this step was to determine whether the model generalized to predict attention phenotypes (*d*’) in an external dataset. The summary model was also applied to ABIDE and to HBN to determine if the model predicted SRS scores in external samples. ABIDE, autism brain imaging data exchange; ADOS, Autism Diagnostic Observation Schedule; CPM, connectome-based predictive model; *d*’, d-prime (attention phenotype in the Adult Attention Sample); HBN, Healthy Brain Network; SRS, social responsiveness scale.

Participants in the Yale youth sample completed three different scanning conditions (Figure 2; see Methods for further description of each task). We note that the SSA clips were counterbalanced across participants; the other scan conditions were not (Supplemental Materials; Supplemental Figure 1A). A standard preprocessing approach was used to generate connectomes^33,63–65^ from the different scanning conditions using a 268-node atlas^66^. For each subject, the mean time-course of each region of interest (“node”) was computed, and the Pearson correlation coefficient was calculated between each pair of nodes to achieve a symmetric 268 x 268 matrix of correlation values representing connections between nodes (“edges”). The Pearson correlation coefficients were then transformed to *z*-scores via a Fisher transformation, and only the upper triangle of the matrix was considered, yielding 35,778 unique edges.

**Figure 2.**
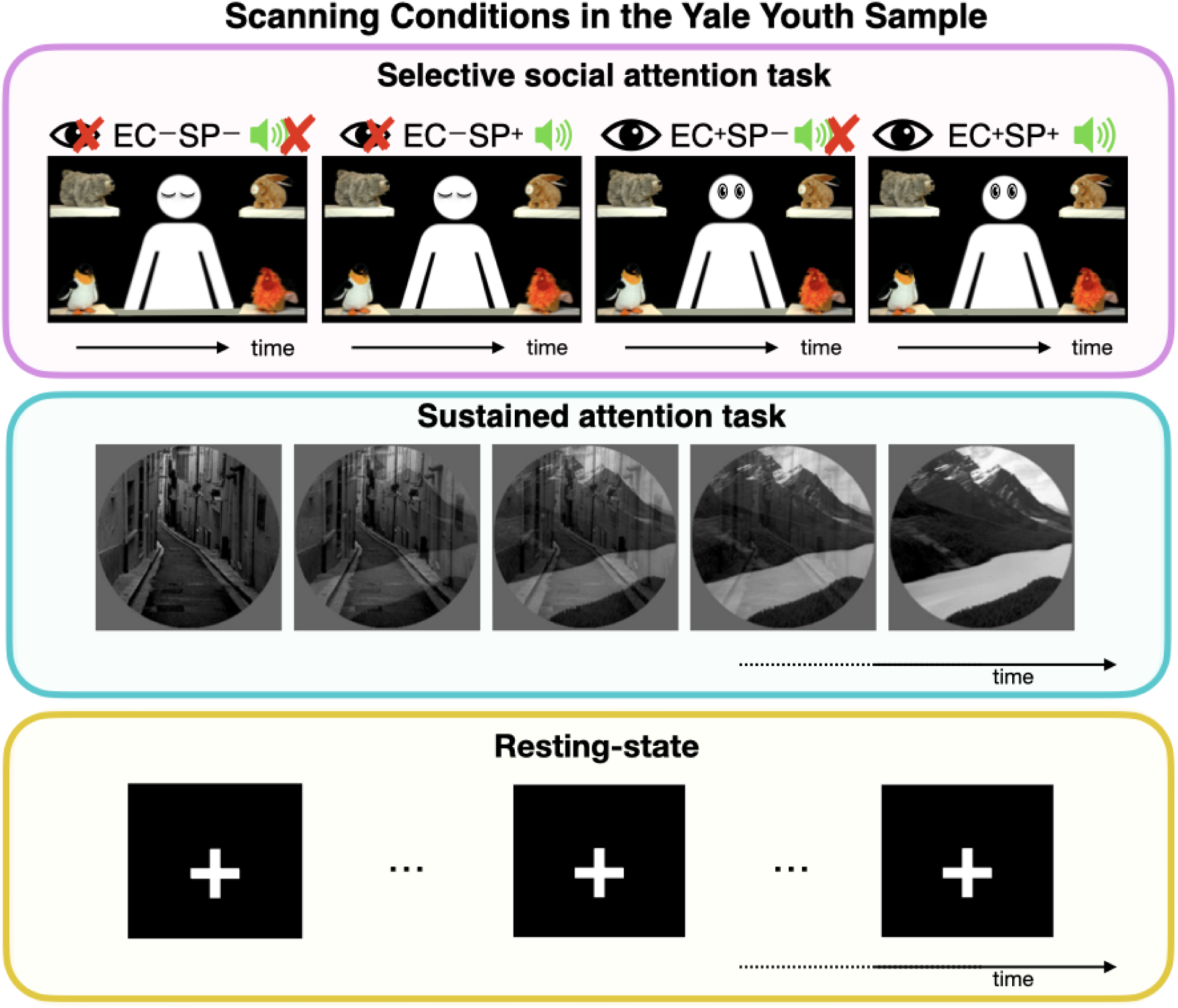
A schematic showing the scanning conditions used in the Yale youth sample. Top panel: free-viewing selective social attention task. Four conditions were shown to participants: no eye contact, no speech (EC-SP-); no eye contact, with speech (EC-SP+); eye contact, with no speech (EC+SP-); and eye contact, with speech (EC+SP+). No responses were required of participants during the different conditions. Middle panel: the gradual onset continuous performance task (gradCPT) was used as a sustained attention task. Grayscale pictures of cities and mountains were presented with images gradually transitioning from one to the next; button presses were required for city scenes and withheld for mountain scenes. Bottom panel: resting-state condition, in which the participants viewed a fixation cross. Please see the Methods section for further details about each scanning condition.

### Prediction performance is highest in the Yale youth sample using task data

We first assessed which scanning condition resulted in the highest prediction performance of autistic traits in the Yale youth sample. To ensure consistent amounts of data across scanning conditions, we discarded frames from the end of gradCPT and resting-state runs, such that the total amount of data was the same as from the Selective Social Attention task runs (four minutes of data). CPM^23^ was then used to assess prediction performance of ADOS scores (Supplemental Figure 1B) and was repeated 500 times. Head motion was controlled for during CPM as before^28,67,68^. The median performing model is represented in the text below, as well as prediction ranges where appropriate; significance was assessed via permutation testing (Methods).

We found differential performance across the various task conditions (Figure 3; Supplemental Table 2). For example, performance using the resting-state data was quite low (rest 1, Spearman’s rho = 0.093, *P*-value = 0.23; rest 2, Spearman’s rho = 0.18, *P*-value = 0.1240), and prediction performance was noted to have substantial variance (i.e., using data from resting-state run 1, the minimum Spearman’s rho =-0.2017, maximum Spearman’s rho = 0.337, with 15% of the prediction performance scores below zero). Performance was also quite low in the SSA condition with no eye contact and no speech (EC-SP-; Spearman’s rho =-0.106, *P*-value = 0.822). Surprisingly, there was large variance in prediction performance scores using the SSA condition with eye contact and speech (EC+SP+; minimum Spearman’s rho =-0.172; maximum Spearman’s rho = 0.323, with 7.2% of the prediction performance scores below zero). Prediction performance was higher in the other SSA conditions, but was not statistically significant after correcting for multiple comparisons (EC+SP-: Spearman’s rho = 0.251, *P*-value = 0.052; EC-SP+: Spearman’s rho = 0.266, *P*-value = 0.034). The only condition that resulted in statistically significant brain-behavior predictions was gradCPT 1 (Spearman’s rho = 0.441, *P*-value = 0.004, corrected). See Supplemental Table 2 for statistics for all CPM analyses.

**Figure 3.**
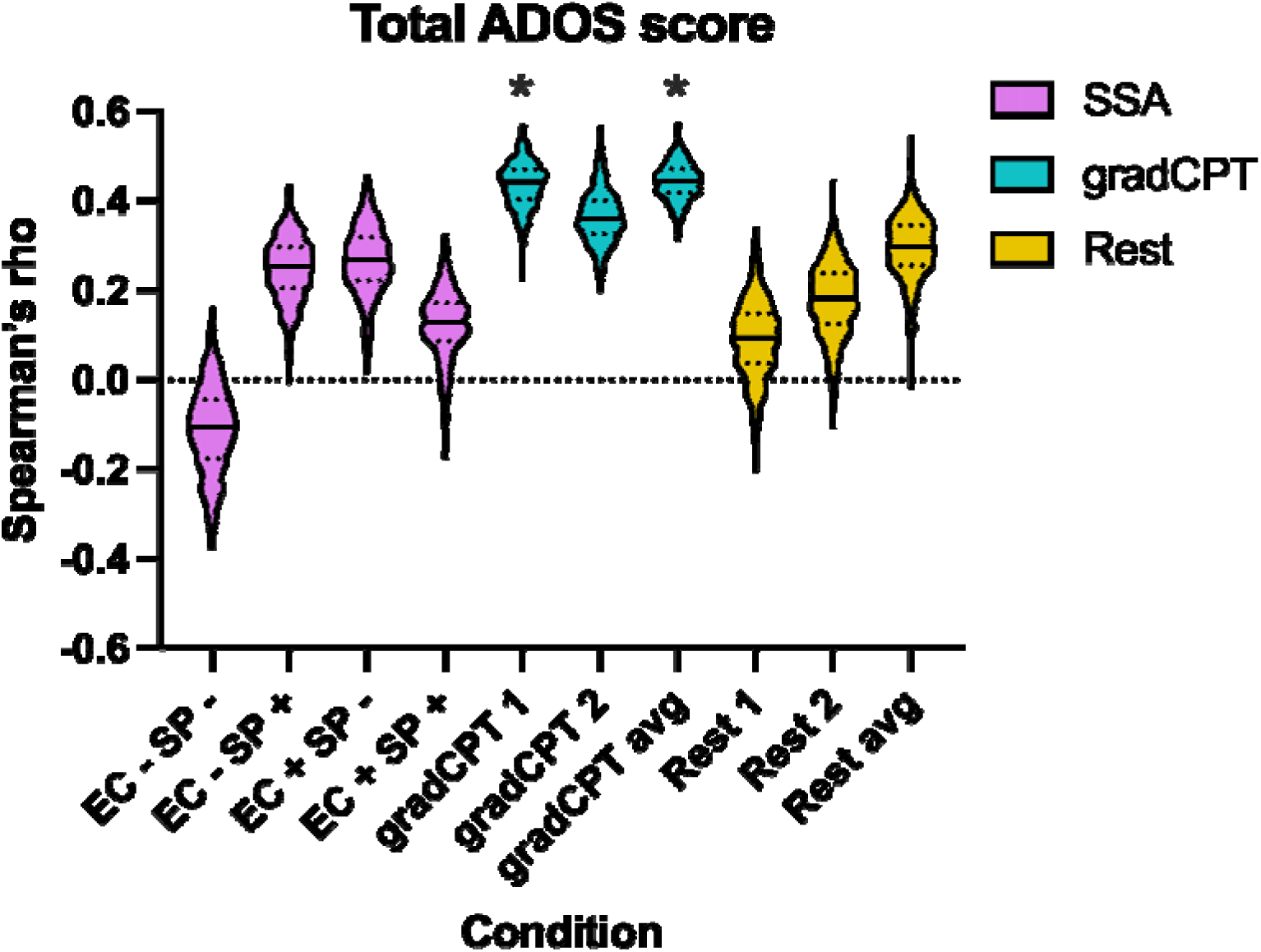
CPM prediction performance across different scanning conditions for total ADOS scores. The scan condition is shown on the x-axis; on the y-axis, Spearman’s rho is shown for the correlation of predicted and actual ADOS scores. For each condition, the median of the 500 iterations is shown as a solid black line in the violin plot; quartiles, as dotted lines. The Selective Social Attention task conditions are shown in purple, gradCPT in turquoise, and resting-state data in yellow. Statistical significance was obtained using permutation testing as described in the Methods. Note that one-side significance testing was used, as we are interested in statistically significant positive predictions only. Asterisk (*) indicates statistical significance after correcting for multiple comparisons using the Benjamini–Hochberg method^135^ (correcting for ten tests: two for gradCPT, four for SSA, two for resting-state, one for gradCPT average, and one for resting-state average). Exact Spearman’s rho and *P*-values for statistically significant conditions, gradCPT1: Spearman’s rho = 0.441, *P*-value = 0.004; gradCPT average: Spearman’s rho = 0.445, *P*-value = 0.002. ADOS, autism diagnostic observation schedule; Avg, average; EC-SP-, no eye contact, no speech; EC-SP+, no eye contact, with speech; EC+SP-, eye contact, with no speech; EC+SP+, eye contact, with speech; mm, millimeters; SSA, selective social attention task.

Prediction performance has been noted to increase with increasing amounts of data^69^, possibly due to an increase in reliability of functional connectivity estimates^70–72^. We tested this possibility by combining data from gradCPT 1 and gradCPT 2, as well as resting-state session 1 and resting-state session 2. More data led to a slight increase in prediction performance using both gradCPT and rest (Figure 3), though only gradCPT prediction performance was statistically significant after multiple comparisons correction (gradCPT average: Spearman’s rho = 0.445, *P*-value = 0.002, corrected; rest average: Spearman’s rho = 0.296, *P*-value = 0.038).

To ensure results were internally consistent, we repeated the CPM analysis using a multiverse approach, which assesses how results are affected by different analytical choices^73^. The point of this approach is not to determine what CPM pipeline results in the highest prediction performance. Instead, the goal is to assess various analytical scenarios and determine how arbitrary modelling choices impact CPM performance. In the Yale youth sample, we first adjusted CPM models for age, sex, and IQ. Encouragingly, we found similar results to above: gradCPT results in the highest prediction performance; the SSA task with no eye contact and no speech results in the lowest (Supplemental Table 2). The other SSA task conditions did not tend to result in high predictions; rest performance was also low. As an additional internal control, we assessed if gradCPT task performance differed among those with autism compared to those without; we observed no difference in performance on the attention task (mean gradCPT score of participants with autism = 2.59 (st. dev. 0.96); mean score of participants without autism = 2.66 (st. dev. 0.86; (*t*(55) = −0.27, *P* = 0.79). Further, we conducted additional controls in which we modelled additional autistic traits with less skewed distributions than the ADOS scores used above; we again found successful prediction of ADOS uncalibrated scores, SRS scores, and ADOS scores when balancing autistic and non-autistic participants (Supplemental Figure 2; Supplemental Table 3).

We continued with the multiverse analysis and repeated CPM using the same pipeline as above, except instead of predicting total ADOS scores, we attempted to predict the social affect and the restricted and repetitive behaviors subscales of the ADOS. We observed the same overall trend—gradCPT tends to result in the highest prediction performance, and the resting-state and SSA task with no eye contact and no speech performed the poorest (Supplemental Figure 1C; Supplemental Table 2). The SSA tasks resulted in increased prediction performance of social affect scores, but predictions were not significant after controlling for multiple comparisons. Prediction performance of restricted and repetitive behaviors using the SSA data tended to be low.

To ensure results were robust to preprocessing pipeline, we reprocessed functional data by altering the functional parcellation (using a 368-node instead of a 268-node atlas); we also repeated analyses without performing global signal regression. In both cases, we observed a similar pattern: gradCPT average had the best absolute prediction performance across pipelines tested, while the SSA and rest conditions tended to vary, with many prediction iterations below zero (Supplemental Figure 1C; Supplemental Table 4). Interestingly, we observed, as have others^33,74,75^, that GSR decreased the strength of brain-behavior relationships, while the overall prediction patterns across scans remained consistent. In sum, these results suggest that the CPM findings observed here were not being driven by a particular processing choice.

Finally, given that we are using a dimensional approach to predict ADOS scores, and ADOS scores are by definition higher in those with autism, we explored if case versus control classification was possible. (Note there is no strict ADOS cut-off to classify autism—it is simply one component of a diagnostic evaluation—but scores tend to be > 5 in those diagnosed.) Modifying CPM and balancing cases and controls, we observed gradCPT data resulted in the highest sensitivity, specificity, and accuracy in case-control classification, particularly the gradCPT average data (Supplemental Figure 1D; Supplemental Table 4). The fact that gradCPT afforded the highest predictions reiterates findings from the dimensional CPM—that task brain states requiring sustained attention tend to outperform those from the SSA and resting-state conditions.

### External validation of predictive models—attention prediction in the adult attention sample

Having determined that data derived from attention tasks are best for predicting autistic traits, we next assessed generalizability of the attention predictive model. Previously, we have shown it is possible to build predictive models of sustained attention in the Yale youth sample and that such a model is related to autistic traits^28^. Therefore, we assessed the extent to which predictive models of autistic traits are related to sustained attention. To ensure generalizability was not driven by sample-specific noise, we tested the predictive model in an external dataset of individuals performing the same gradCPT task (n=25 neurotypical adults, 13 females, mean age = 22.8 years, st. dev. = 3.5 years)^36^. Hereafter, we refer to this dataset as the “adult attention sample” (Figure 1). The behavioral outcome of interest in this sample is performance on the gradCPT, *d’* (sensitivity), the participant’s hit rate minus false alarm rate (mean *d*’ = 2.11, st. dev. = 0.92).

We determined which edges tended to contribute consistently to successful prediction of ADOS phenotypes in the Yale youth sample (see Methods, ‘Testing generalizability of the ADOS network’) using the model generated from average gradCPT data in the prediction of total ADOS scores. The resulting model (the ‘ADOS consensus network’) was used to determine if there was a relationship between predicted ADOS scores and *d*’ scores in the adult attention sample. Specifically, we used the fMRI gradCPT task data from the adult attention sample to generate a predicted ADOS score. Predicted ADOS scores were then compared to actual *d*’ scores across participants to assess accuracy. We point out this differs from the Yale youth sample, where we were able to compare predicted ADOS scores with observed ADOS scores. In the adult attention sample, the goal was to assess the relationship between the model (trained to predict ADOS) and attention (*d*’).

We observed a statistically significant relationship between predicted ADOS scores and *d*’ scores (Spearman’s rho =-0.56, *P* = 0.0049, corrected; Figure 4A). Specifically, higher predicted ADOS scores were associated with lower *d*’ scores, indicating poorer performance on the task and implying lower sustained attention. To ensure results were robust, we repeated analyses controlling for potential confounds; predictions remained high when adjusting for participant head motion (Spearman’s rho =-0.56, *P* = 0.0043), participant sex (Spearman’s rho =-0.49, *P* = 0.0164), and participant age (Spearman’s rho =-0.55, *P* = 0.0066). In addition, we also assessed the relationship between predicted ADOS scores and *d*’ scores using only the ADOS positive network and then only the ADOS negative network. We observed a statistically significant negative correlation in the ADOS positive model (Spearman’s rho =-0.59, *P* = 0.0021, corrected; Figure 4B) and in the ADOS negative model (Spearman’s rho =-0.46, *P* = 0.023, corrected; Figure 4C).

**Figure 4.**
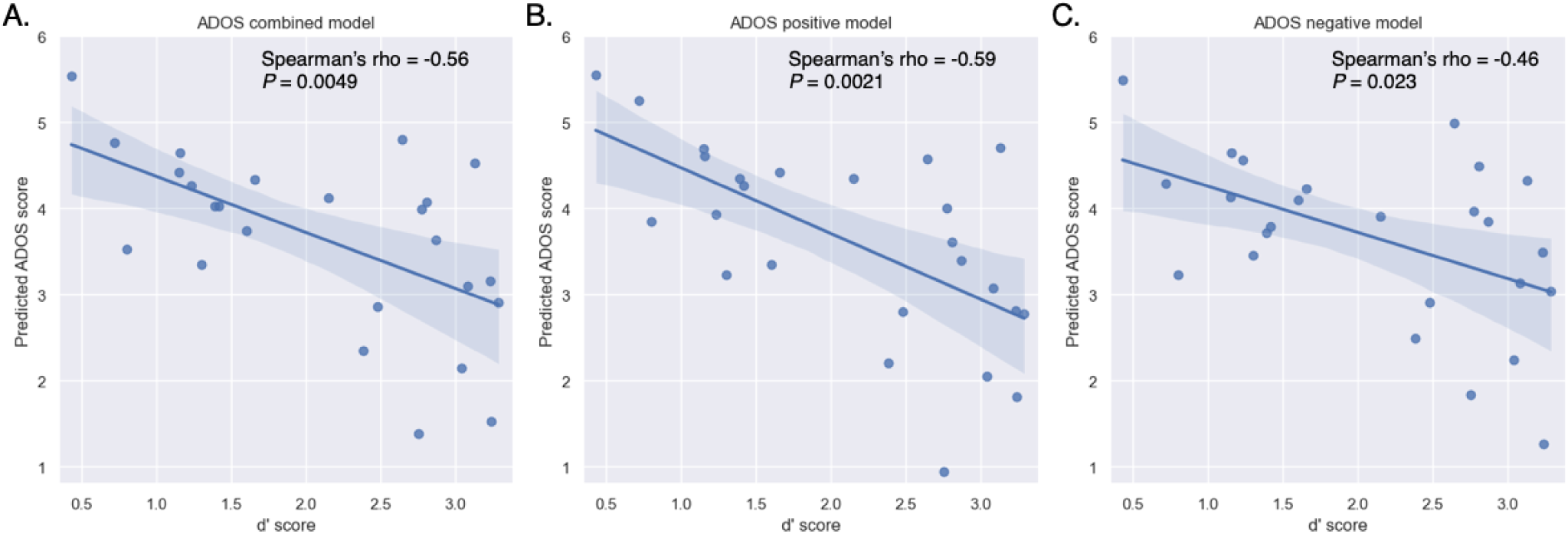
Generalization of the ADOS consensus network in the adult attention sample. A. Results using the combined network model. B. Results using the positive-association network model. C. Results using the negative-association network model. Predicted ADOS scores are indicated on the *y*-axis; *d*’ scores, on the *x*-axis. Higher predicted ADOS scores are associated with lower *d*’ scores, indicating poorer performance on the task and implying lower sustained attention. Data are presented as a regression line (solid line) +/- 95% confidence intervals (shaded areas). Spearman’s rho was calculated and was used to assess statistical significance (two-sided). The Benjamini–Hochberg method^135^ was used to correct for three tests. ADOS, autism diagnostic observation schedule; *P* = *P*-value.

Lastly, we altered the stringency of how often an edge had to be included in the ADOS consensus model (Methods). We observed consistent results across a range of thresholds (Supplemental Table 5), increasing confidence that there is a relationship between ADOS network strength and *d*’ scores. In sum, these results suggest that the predictive model of autistic traits captures variance related to sustained attention.

### External validation of predictive models—social responsiveness prediction in ABIDE and HBN

After finding we could successfully predict attention scores, we set out to determine if the predictive model from the Yale youth sample generalized to predict social responsiveness in a large sample of participants from ABIDE (n=229, 65 females; mean age = 10.45 years, st. dev. = 1.8 years; mean IQ = 113.7, st. dev. = 15.1; 77 individuals with autism)^76,77^ described elsewhere^24^. We used the same approach as in the adult attention sample to assess generalizability. Specifically, we used the resting-state data from ABIDE and applied the ADOS consensus model to predict social responsiveness scale (SRS) scores^78^ across participants (Methods). As with the other test of generalizability above, predicted ADOS scores were then compared to actual SRS scores to assess accuracy.

We observed successful prediction of all SRS scales tested (Figure 5). In particular, the model generalized to predict SRS total scores (Spearman’s rho = 0.17, *P* = 0.008, corrected, Figure 5A), as well as SRS subscales quantifying communication (Spearman’s rho = 0.15, *P* = 0.028, corrected, Figure 5B), mannerisms (Spearman’s rho = 0.21, *P* = 0.001, corrected, Figure 5C), and motivation (Spearman’s rho = 0.16, *P* = 0.016, corrected, Figure 5D). We also tested prediction of each SRS scale after adjusting for participant age, sex, and head motion; predictions were essentially unchanged, further supporting that the ADOS model is capturing variance related to the SRS scales (Supplemental Table 6). As above, we altered how often an edge had to be included in the ADOS CPM and retested predictions. In every case, we observed similar predictions across various thresholds for all SRS scales (Supplemental Table 6).

**Figure 5.**
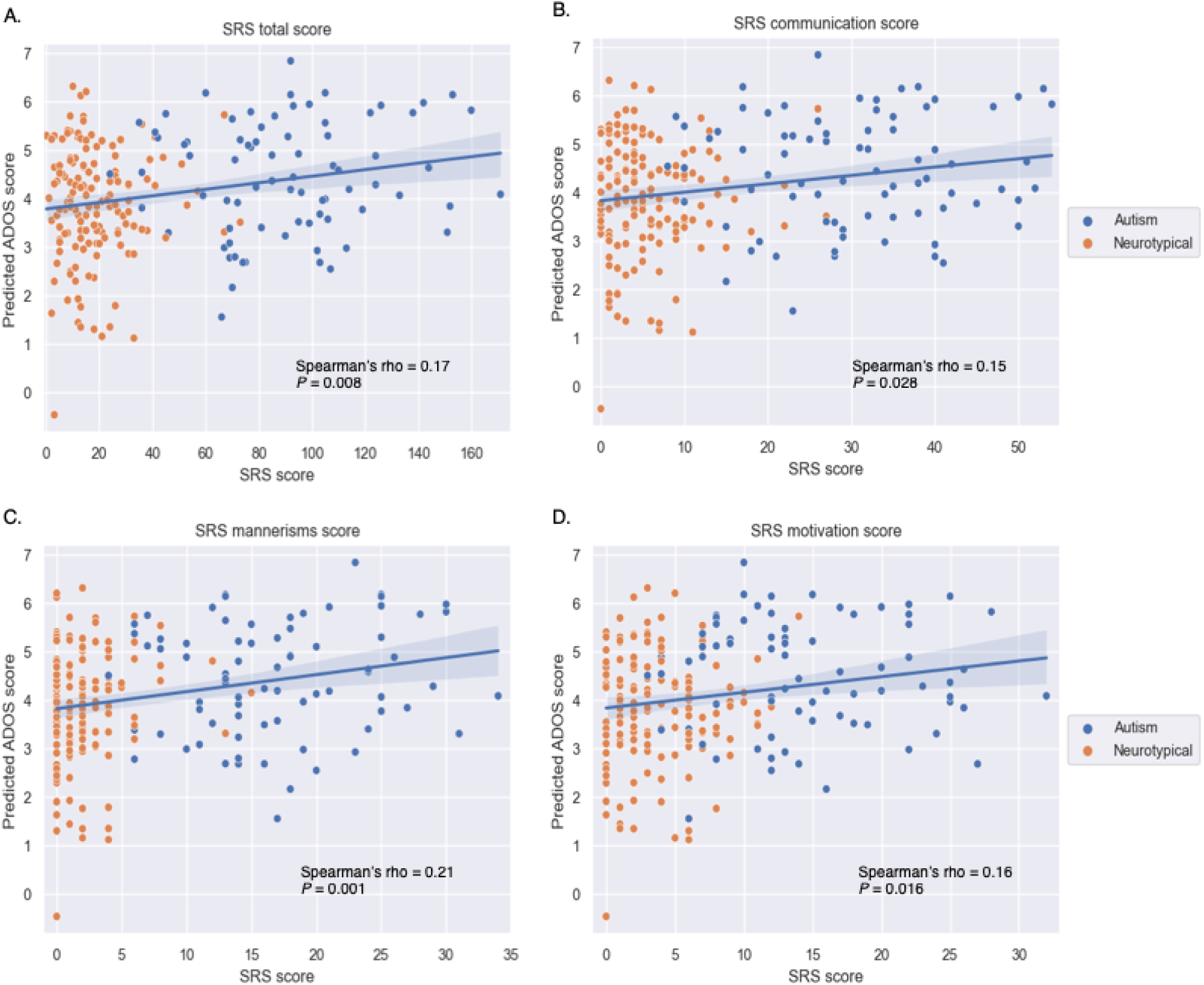
Generalization of the ADOS consensus network to ABIDE. A. SRS total score results. B. SRS communication score results. C. SRS mannerism score results. D. SRS motivation score results. For all plots, actual SRS scores from each subscale are indicated on the *x*-axis; predicted scores, on the *y*-axis. Data are presented as a regression line (solid line) +/-95% confidence intervals (shaded areas). Spearman’s rho was calculated and was used to assess statistical significance (two-sided). The Benjamini–Hochberg method^135^ was used to correct for four tests. ADOS, autism diagnostic observation schedule; *P* = *P*-value.

We performed a similar test of generalization using data from the Healthy Brain Network (HBN) (n=643, 264 females; mean age = 11.01 years, st. dev. = 2.63 years; mean IQ = 101.89, st. dev. = 16.7; 107 participants had a diagnosis of autism; 302 had a diagnosis of ADHD). Using functional data obtained while participants watched a naturalistic movie clip (scenes from Despicable Me; Methods), we applied the ADOS consensus model to predict social responsiveness scale (SRS total) T-scores (mean = 56.36, st. dev. = 11.21)^78^ across participants. Predicted ADOS scores were then compared to actual SRS scores to assess accuracy. Controlling for head motion, we again observed the model generalized to predict SRS scores (Spearman’s rho = 0.1002, *P* = 0.0111). Like before, we tested a variety of summary models of differing sizes. We observed that results were stable when the consensus model had both more and fewer edges, supporting that our results were robust to model size (Supplemental Table 7). Taken together, these data indicate the ADOS model from the Yale youth sample generalized to predict aspects of sociality in ABIDE and HBN.

### Neuroanatomy of predictive edges

We next visualized brain connections in the ADOS consensus model. The network comprised 2,014 total edges (1,001 edges in the positive-association and 1,013 edges in the negative-association network), approximately 5.6% of the connectome, in line with other models that have generalized to predict autism and attention symptoms^24,28^. Edges across the brain were represented in the model, constituting a complex, distributed network (Figure 6A-B). In particular, connections within and between heteromodal association networks were found to contain the highest fraction of edges (Figure 6C-D; note that results have been corrected for differing network size)^28^. For instance, the top three network pairs containing the greatest proportion of edges in the positive-association network involved medial frontal, frontoparietal, or default mode networks. In the negative-association network, the top three network pairs involved connections within and between medial frontal, frontoparietal, or default mode networks (e.g., in this case, the top network pair comprised connections within the medial frontal network; the next highest network comprised connections between the medial frontal and frontoparietal networks; and the third highest network connected the medial frontal and default mode networks). In addition, 704/1001 of the edges in the positive-association network and 535/1013 of the edges connected to medial frontal, frontoparietal, or default mode networks. We performed further visualizations using slightly different thresholding techniques; these analyses again showed that association networks were important in the ADOS consensus model (Supplemental Figure 1E).

**Figure 6.**
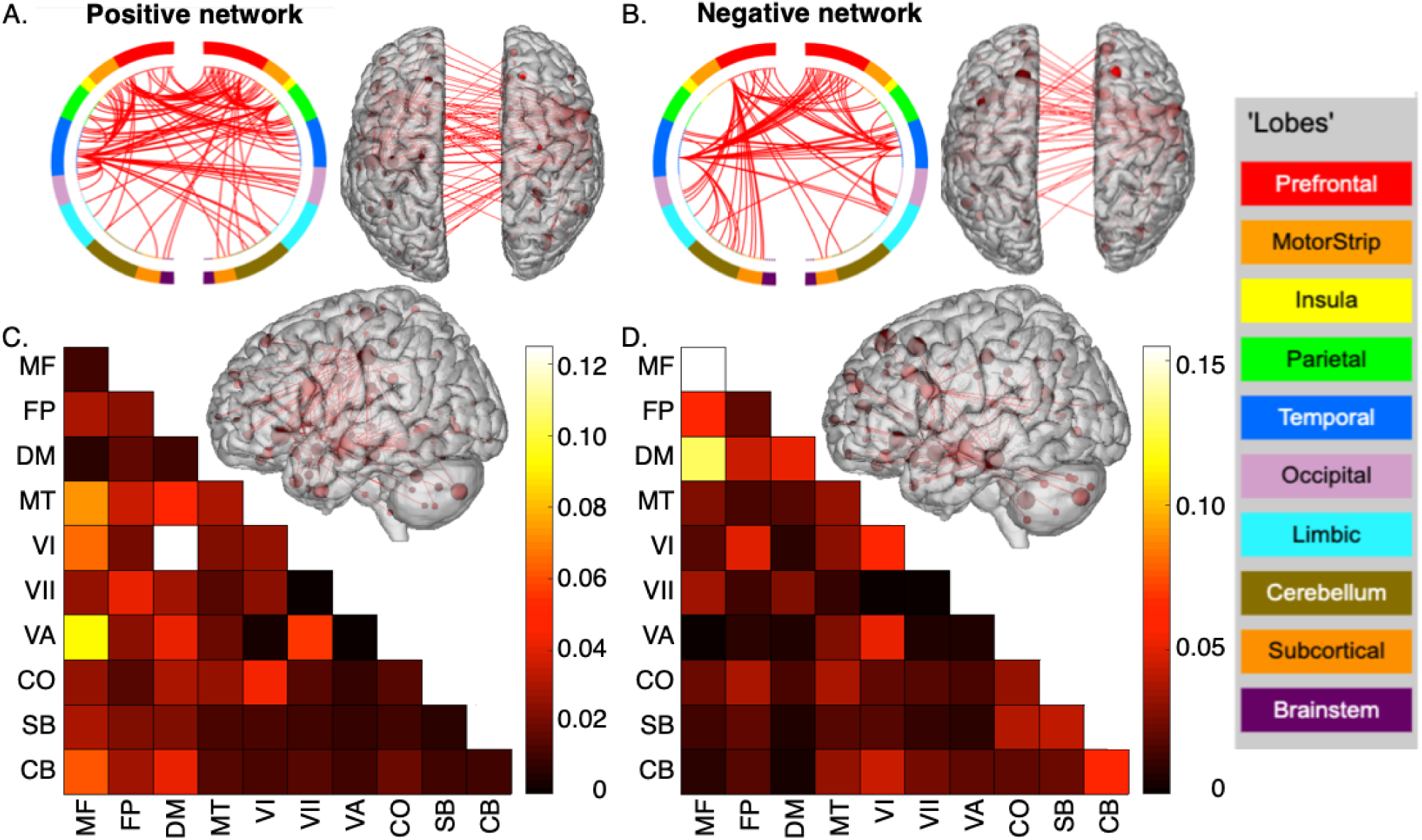
Neuroanatomy of ADOS consensus network. A. The positive-association network. B. The negative-association network. For both A and B: a circle plot is shown in the upper left. The top of the circle represents anterior; the bottom, posterior. The left half of the circle plot corresponds to the left hemisphere of the brain. A legend indicating the approximate anatomic ‘lobe’ is shown to the left. The same edges are plotted in the glass brains as lines connecting different nodes (red circles); in these visualizations, nodes are sized according to degree, the number of edges connected to that node. To aid in visualization, we have thresholded the matrices to only show nodes with a degree threshold > 25. C. Matrix of the positive-association network. D. Matrix of the negative-association network. For both C and D: The proportion of edges in a given network pair is shown; data have been corrected to account for differing network size. MF, medial frontal; FP, frontoparietal; DM, default mode; MT, motor; VI, visual I; VII, visual II; VA, visual association; CO, cingulo-opercular; SB, subcortical; CB, cerebellum.

### Testing for convergence of task versus rest CPM findings

As a last test of generalizability, we repeated analyses, except we reversed the order of which dataset was used to build the original model. As our primary aim was determining if task data outperformed resting-state data in the prediction of autistic traits, and ABIDE does not have task data, we used the Healthy Brain Network (HBN) sample. Specifically, we used data from two conditions in which participants watched movies (‘Despicable Me’ and ‘The Present’) in the scanner, along with a resting-state condition. Although HBN does not contain the exact same tasks as the Yale youth sample, because the movie watching conditions comprise a continuous task requiring sustained attention to an evolving narrative involving multiple characters interacting in a complex scene, the sample still allows us to test broadly if there is a relationship between autistic traits and attention. Note that the naturalistic movies stand in distinction to the SSA tasks, which were designed to include only one actress speaking at a time in short phrases (i.e., no narrative content or social interactions are otherwise presented by the participant during the SSA tasks).

After ensuring all scan conditions had the same amount of data per scan (Methods), we repeated CPM, controlling for participant head-motion as above, and attempted to predict SRS scores. We observed successful prediction in the movie-watching conditions (median performing model from Despicable Me: Spearman’s rho = 0.14, *P*-value < 0.001, corrected; median performing model from The Present: Spearman’s rho = 0.11, *P*-value < 0.0001, corrected). Prediction using resting-state data was non-significant (median performing model: Spearman’s rho = 0.05, *P*-value = 0.102, corrected (Supplemental Figure 1F).

To assess generalizability of the model, we used connections that tended to contribute to successful predictions in the HBN DM condition to generate a consensus model (Methods). The resulting DM consensus model was used to assess if there was a relationship between predicted SRS scores and ADOS scores in the Yale youth sample. Specifically, we used the gradCPT data in the Yale youth sample and applied the DM consensus model to predict social responsiveness scale (SRS) scores^78^ across participants (Methods), again controlling for head motion. Predicted SRS scores were then compared to actual ADOS scores to assess accuracy. Successful prediction was achieved (Spearman’s rho = 0.26; *P*-value = 0.0422). To ensure results were robust, we repeated analyses using different thresholds when generating the consensus mask; we found results were stable across both looser and stricter thresholds (Supplemental Table 8). In sum, the analyses in this section replicate our initial finding that task data outperform resting state data in predicting autistic traits and that such a model can generalize in external samples.

## Discussion

We determined that using functional connectivity calculated from data acquired during gradCPT resulted in the prediction of autistic traits. The resulting model generalized to independent samples to predict attention and social phenotypes in neurotypical participants and those with autism. Altogether, results highlight the potential of using in-scanner tasks, particularly those demanding sustained attention, to more accurately determine brain-behavior relationships in clinical samples.

There is a rich history of using tasks to probe the cognitive architecture of autism^79–84^. Nevertheless, most fMRI brain-behavior prediction studies in autism that use machine learning techniques have typically relied on resting-state data (see^4^ for a recent review). Our results suggest that by optimizing the brain state under which data are acquired through task engagement^85^, more accurate brain-behavior relationships can be studied^29^. Improved brain-behavior mapping increases the potential clinical utility of neuroimaging approaches^86^ and might help obtain a more accurate picture of brain circuits underlying the complex phenotypic landscape of autism. Tasks also offer the advantage of improving the reliability of task-engaged functional connections^87^. More generally, the results obtained here are in line with other work in neurotypical populations, indicating that predictions of phenotypes improve when using task as opposed to resting-state data^29,33,34,36,41,88,89^. We note that resting-state studies still retain utility, particularly in terms of ease of data collection and their ability to facilitate the collation of large datasets across centers.

Despite tasks outperforming rest-based models, our data indicate that not all tasks are equal. Our original hypothesis regarding prediction performance was that, reflective of social difficulties in autism, the SSA tasks would result in the highest prediction of ADOS scores. That an attention task—superficially unrelated to the core aspects of autism—outperformed a task designed to probe social abilities reinforces the importance of empirical work. The current results suggest that simply because a scanning condition is ostensibly related to a given phenotype does not mean that a brain-behavior relationship can be found in neurodevelopmental conditions.

It is perhaps puzzling that the SSA clips did not result in higher prediction performance.

This could be due to the passive nature of the SSA clips, allowing the participants to let their minds wander^55,56^. As such, SSA clips could be interpreted as resembling resting-state^112^. The unconstrained nature of the resting-state might be suboptimal for probing certain aspects of brain-behavior relationships^29^. In parallel, it is perhaps also surprising that the gradCPT data resulted in the highest prediction performance. Beyond attention, the highly structured, rules-based design of gradCPT may effectively highlight variations in networks linked to autistic traits given the rules-based tendencies observed in autism^113^.

Regardless of the exact mechanism, our finding that gradCPT led to the prediction of autistic and attention phenotypes adds to the growing literature suggesting an important link between autism and attention at the neurobiological level. Previous studies^24,47,93,94^ have indicated that complex models spanning numerous functional networks are important for attention in autism. In addition, the size of the ADOS consensus model is of interest (approximately 5% of the connectome) and is in line with the size of other models that have generalized to predict autism and attention symptoms^24,28^. Notably, these models tend to share a few characteristics with the ADOS consensus model. Edges tend to originate across the entire brain, linking within and between most canonical network pairs. This finding underscores that there is no one region or network preferentially involved in a given phenotype, consistent with other psychiatric conditions^95^. The fact that distributed brain connections mediate complex behaviors is a theme observed in multiple species, including the global brain dynamics underlying homeostatic sleep behaviors in *Caenorhabditis elegans*^96^, to long-range connections underpinning social decisions in primates^97^.

While the ADOS model is complex, post-hoc analyses revealed other neurobiological trends. Similar to the ADOS model, previous models involved in predicting attention and autism all tend to have particular representation in heteromodal association cortices^67^, including the default mode (recently reviewed in^98^). In work involving CPM, the default mode network tends to be important, especially in the negative networks^24,26,28^. In CPM, edges comprising the negative network are selected based on a negative correlation with the behavior of interest. In the present work, it follows that higher connectivity in the default mode, typically associated with more mind-wandering or internally-directed states^99^, would be associated with lower attention abilities, as well as potentially higher autistic traits. (A recent mega-analysis has observed hyperconnectivity involving the DMN in both autistic and ADHD participants as well.^100^) It is plausible that interactions among association networks—including not only the default mode, but also the medial frontal and frontal parietal networks—are needed to coordinate the complex cognitive demands associated with higher-level brain functions^101^, such as sustaining attention and picking up on social cues.

We contend that autistic traits and attentional abilities are hence multifaceted, interrelated processes. Because the CPM also predicts attention in the adult sample, it indicates that the attention task state is appropriately influencing brain circuits of interest involved in modulating attention in the Yale autism sample. That the CPM goes on to predict a different phenotype related to a core component of autism—namely, social difficulties—reiterates the shared relationships between autistic features and attention. Indeed, autism and ADHD symptoms have long been acknowledged to overlap^49–51,104^. In a more extreme example, it is possible the CPM is more related to a broader pathology and could be applicable, to some degree, to many other phenotypes, similar to the p-factor^105^. Instead of commenting on the validity of the p-factor as a single unifying construct^106^, we simply note that there are numerous examples of redundancy^107^ and degeneracy in biological circuits^108,109^, especially in brain networks. Further, we do not mean to argue that we have discovered a ‘core autism/attention network.’ Rather, it is logical that these phenotypes, both defined to some extent by how complex information is perceived from the outside world, are associated with overlapping circuitry.

A few additional items warrant discussion in the final paragraphs. Participant IQ in the Yale youth sample and ABIDE was fairly high. More research should be conducted using participants with a broad range of IQ scores to determine which scanning conditions are optimal for prediction performance. The current work relied on a sample of 20/63 participants with autism. Future work could aim to repeat analyses in more participants with autism. The autism dataset contained mainly males; the importance of sex-based differences in brain circuitry^114–116^ and behavioral phenotypes^117^ relevant for autism is increasingly well-described. It is possible that a more active social task requiring participant engagement (unlike the SSA task) could increase prediction performance. Likewise, given the success of the movie-watching conditions of the HBN, perhaps more naturalistic movie clips (demanding the processing of narrative content and complex, interpersonal relationships) are another promising path forward in autism research^118,119^. Further, the role of arousal and its impact on predictive modelling could also be studied. In addition, while we have taken numerous steps to lessen the effect of head motion, it can still impact brain-behavior relationships; this must be kept in mind when interpreting the current work. A potential limitation of the adult attention sample is that it comprises older participants; another possible interpretation is that it strengthens the generalizability of the CPM. That is, the fact the model generalized in a different age range eliminates the potential confound that the model is confounded by age in the Yale youth sample (which in turn would drive successful generalization in a test sample of similar ranges, but would fail to generalize in a sample comprising different ages).

Also, the current work focused on prediction of traits in an adolescent dataset that is relatively small; the adult attention sample was also quite small. Though replicable and generalizable findings can still be determined by using robust methods^120–122^ and we were well-powered in the current study (see Methods for a power analysis), future studies in larger samples are needed. Collecting small, unique samples also facilitates testing across diverse experimental conditions, thereby enhancing generalizability^123^. We contend that in the age of big data, it is essential to continue exploring brain-behavior associations in samples that might not contain thousands of subjects, but comprise unique scanning conditions. Such an approach allows the field to better determine which scans to include in big data endeavors and facilitates the exploration of questions that may be difficult to address in large-scale studies. Balancing the need for large samples with adequate scans per subject is a key question that has received much attention in the neurotypical brain-wide association literature^124,125^. Related studies could be pursued in the autism field. Including participants with moderate autism symptoms to tease apart the brain correlates of more subtle phenotypes will be an important aspect of future work, regardless of exact study design. More studies could also be conducted to assess task design and prediction performance in much younger samples, such as toddlers and young children. Such efforts aim to optimize the detection of brain-behavior relationships at earlier developmental stages, ultimately providing better support for individuals with autism and their families.

## Conclusions

We have shown in a preliminary study that sustained attention tasks, such as gradCPT, can enhance the prediction of autistic traits. Such an approach leads to a robust marker that generalizes to predict attention and social phenotypes in independent samples. Our findings highlight the need to further investigate optimal brain states for modeling phenotypes in autism and related conditions.

## Methods

### Description of datasets

We used four datasets in this work (Figure 1). The first dataset, the Yale youth sample, comprised 63 subjects from a sample described elsewhere (mean age = 11.7 years, st. dev. = 2.8 years; 29 females; mean IQ = 107.8, st. dev. = 15.1)^28,60^. Twenty of the participants had autism (five of whom had ADHD); 11 other participants had a neurodevelopmental condition (five with ADHD, two with anxiety disorder, and four were classified as belonging to the broader autism phenotype)^61^. Participants were scanned on a 3T Siemens Prisma System. See Supplementary Material for full exclusion criteria and imaging parameters. Autism symptoms were scored using the Autism Diagnostic Observation Schedule-2 (ADOS-2)^62^ and were ascertained by trained clinical psychologists; calibrated severity scores were used in the present work for the social affect subscale (mean = 3.2, st. dev. = 2.9), the restricted and repetitive behavior subscale (mean = 4.0, st. dev. 3.3), and the ADOS total score (mean = 3.1, st. dev. = 3.1). This sample was used to conduct CPM, compare how scanning condition impacted performance, and generate a consensus model. Note that although the sample is small, we were well-powered to detect effects. Using the R package “pwr”, with a sample size of 63 subjects, and assuming a significance level of 0.05 (two-sided), a correlation between predicted and observed ADOS scores in gradCPT average of 0.445 corresponds to a power of 0.96.

A second dataset of neurotypical adults, the adult attention sample (n=25, 13 females, mean age = 22.8 years, st. dev. = 3.5 years) was used as a validation dataset and is described elsewhere^36^. Participants were scanned on a 3T Siemens Trio TIM system. This sample was used to determine if the consensus model generalized to predict attention.

A third dataset of individuals with and without autism (n=229, 65 females; mean age = 10.45 years, st. dev. = 1.8 years; mean IQ = 113.7, st. dev. = 15.1) comprising data from ABIDE^76,77^ was used as an additional validation dataset; processing of these data is described elsewhere^24^. Note that 77/229 participants were diagnosed with autism. SRS^78^ raw scores were used from ABIDE and included the following scales: SRS total scores (mean = 42.4, st. dev. = 40.2); SRS communication (mean = 13.8, st. dev. = 14.2); SRS motivation (mean = 7.3, st. dev. = 6.9); SRS mannerisms (mean = 7.1, st. dev. = 8.5). Seventy-seven of the participants had autism. SRS was chosen due to the low numbers of subjects with ADOS scores (when using the exclusion criteria described in^24^), along with the additional quality control exclusion criteria we performed (i.e., there were 229 subjects with SRS data, compared to only 33 with ADOS; see Supplemental Methods). This sample was used to determine if the consensus model generalized to predict SRS scores.

A fourth dataset of individuals (n=643, 264 females; mean age = 11.01 years, st. dev. = 2.63 years; mean IQ = 101.89, st. dev. = 16.7) comprising data from the transdiagnostic HBN sample^126^ was used as an additional validation dataset. In the current study, 107 had a diagnosis of autism, while 302 had a diagnosis of ADHD. Processing of these data is described elsewhere^127^. SRS^78^ total T-scores were used from HBN (mean = 56.36, st. dev. = 11.21). This sample was used to determine if the consensus model generalized to predict SRS scores.

For the Yale youth sample and the adult attention sample, the Yale University institutional review board oversaw and approved the ethics of the study. For HBN, the study was approved by the Chesapeake Institutional Review Board. The data for ABIDE were approved by each of the 16 sites contributing data (i.e. approval was required by each of the home institutions prior to submission)^76^. Where appropriate, informed consent was obtained from the parents or guardians of participants, and participants were paid for their involvement. Written assent was obtained from children aged 13–17 years; verbal assent was obtained from participants under the age of 13 years.

### Preprocessing of functional imaging data

A standard preprocessing approach was used that has been described elsewhere^33,63–65^. Preprocessing steps were performed using BioImage Suite^128^ unless otherwise noted, and included: skull stripping the 3D magnetization prepared rapid gradient echo images using optiBET^129^ and performing linear and non-linear transformations to warp a 268-node functional atlas^130^ from Montreal Neurological Institute space to single subject space. Functional images were motion-corrected using SPM8 (https://www.fil.ion.ucl.ac.uk/spm/software/spm8/).

Covariates of no interest were regressed from the data, including linear, quadratic, and cubic drift, a 24-parameter model of motion^131^, mean cerebrospinal fluid signal, mean white matter signal, and the global signal. Data were temporally smoothed with a zero-mean unit-variance low-pass Gaussian filter (approximate cutoff frequency of 0.12 Hz). Visual inspections were performed after skull-stripping, non-linear, and linear registrations to ensure there were no errors in processing. Head motion was computed as the mean frame-to-frame displacement (FFD) of the participant’s head^60^ (see Supplemental Methods for additional motion control considerations, as well as Supplemental Figure 1G). To ensure consistent amounts of data across scanning conditions, we discarded frames from the end of the gradCPT and resting-state runs, such that the total amount of data was the same as from the SSA task runs (four minutes of data).

Connectomes were generated using a 268-node atlas^22^. For each subject, the mean time-course of each region of interest (“node”) was computed, and the Pearson correlation coefficient was computed between each pair of nodes to achieve a symmetric 268 x 268 matrix of correlation values representing “edges” (connections between nodes). The Pearson correlation coefficients were then transformed to *z*-scores via a Fisher transformation, and only the upper triangle of the matrix was considered, yielding 35,778 unique edges.

The 268-node atlas was chosen because it is a full brain parcellation, covering cortical, subcortical, and cerebellar regions, as well as striking a good balance in terms of number of nodes and reducing computational resources while approximating biologically plausible brain regions^66^. For example, the 268-node atlas results in 35,778 unique edges (because number of edges = ((number of nodes x number of nodes) – number of nodes) / 2). Increasing the atlas size by 100 to 368 nodes results in 67,528 edges, almost doubling the number of edges and thereby doubling compute time. In addition, from a practical perspective, the 268-node parcellation allows generalization testing in the adult attention, ABIDE samples, and HBN samples, which had previously been processed using the 268-node atlas^24,28,36,127^. To ensure results were not being driven by this specific parcellation, we repeated analyses using a 368-node atlas^132^ (which was generated using a different approach, namely, a spectral clustering algorithm in the cortex, anatomical labelling of structures in subcortical regions, and by incorporating cerebellar regions defined elsewhere^133^). We found CPM results did not differ, consistent with other brain-behavior modelling studies showing that results tend to be stable across a range of parcellations^33,74^.

### Scanning conditions in the Yale Youth Sample

1) *Scanning condition one: the free-viewing selective social attention task*

Participants completed a novel version of a free-viewing selective social attention (SSA) task^55,58^ in which an actress is presented at the center of the screen and is surrounded by objects in corners of the screen (Figure 2). Four types of clips were used in which the presence of speech (SP) and eye contact (EC) were manipulated. The first condition included clips in which the person smiled and made eye contact with the camera while speaking in full sentences (e.g., “Have you ever seen a monkey? Monkeys eat bananas, swing in trees, and chase each other.”; this was designated as the EC+SP+ condition). The second condition included a direct gaze condition with no speech (EC+SP-), in which the person smiled directly at the viewer while remaining silent. The third condition consisted of the person looking down at the table while speaking in full sentences (EC-SP+). The fourth condition consisted of the person looking down at the table and not speaking (EC-SP-). Each clip lasted two minutes and was shown twice over four runs, such that eight clips were shown in total. To allow successful scene transitions in between sentences, the direct gaze and speech condition lasted 2 minutes and 8 seconds. The speech with no eye contact condition lasted 2 minutes and 6 seconds. In between clips during each run, a white fixation cross on a black background was shown for 15 seconds. Clip order was counterbalanced across participants (see Supplemental Materials for more about the counterbalancing of clips, as well as study design considerations of the Yale youth sample). Clip conditions were concatenated across runs, such that each resulting connectivity matrix comprised four minutes of data from a single scanning condition. Both gradCPT and the SSA task were presented using Psychtoolbox (version: 3.0.14; http://psychtoolbox.org/; MATLAB version R2018a) on a Lenovo IdeaPad 720S computer, with Ubuntu 16.04 LTS installed.

2) *Scanning condition two: the gradual onset continuous performance task (gradCPT)*

The gradual onset continuous performance task (gradCPT; Figure 2) ^36,53,54^ was used in datasets one and two. The gradCPT tests sustained attention and inhibition, producing a range of performance scores across neurotypical^53,54^ and neurodiverse populations^28^. Participants viewed grayscale pictures of cities and mountains presented at the center of the screen, with images gradually transitioning from one to the next every 1,000 ms. Subjects were instructed to respond with a button press for city scenes and to withhold button presses for mountain scenes. City scenes occurred randomly 90% of the time. Performance was calculated using *d’* (sensitivity), the participant’s hit rate minus false alarm rate. The task took 5 minutes to complete; participants completed the task twice. Note that because of differences in task timing between gradCPT, the selective social attention task, and resting-state, we trimmed the gradCPT and resting-state data to include 4 minutes of data per scan (to match the selective social task time length of 4 minutes).

Subjects in the adult attention sample also performed gradCPT; the same parameters were used as above, except scene transitions took 800 ms.

3) *Scanning condition three: resting-state data*

Resting-state data were also obtained. Subjects were instructed to keep their eyes open, relax, and think of nothing in particular while they viewed a white fixation cross on a black screen. Each scan lasted five minutes and was repeated twice per participant. Resting-state data were also obtained in the ABIDE sample^76,77^.

### Scanning conditions in the HBN sample

We considered three conditions when performing CPM. Specifically, we used functional data from two separate naturalistic movie conditions, in which participants watched *Despicable Me* and *The Present*, as well as a single run of resting-state data. Because of differences in scan time across conditions, we truncated time courses so they were consistent across all three conditions, so that all matrices were generated using four minutes of data (the length of *The Present*).

### Connectome-based predictive modelling

CPM^23^ (Supplemental Figure 1B) was used to predict ADOS scores from functional connectivity data in the Yale youth sample. Briefly, using 10-fold cross-validation, connectivity matrices from a given scan condition and ADOS scores were split into an independent training set including subjects from 9 folds and a test set including the left-out fold. Linear regression was used to relate edge strength to ADOS score in the training set. Edges most strongly associated with ADOS scores were selected (feature selection threshold of *P* = 0.05) for both a ‘positive network’ (in which increased connectivity was associated with higher ADOS scores) and a ‘negative network’ (in which increased connectivity was associated with lower ADOS scores). We used partial correlation to control for mean participant head motion at the feature selection step^28,67,68^. Note that this is the ‘base model’ we are considering throughout the results (i.e., the results shown in Figure 3 used partial correlation to control for motion). Mean network strength was computed in both the positive and negative networks, and the difference between these network strengths was computed (‘combined network strength’), as in previous work ^33^.

A linear model was then calculated relating combined network strength to ADOS scores in the training set. In the last step, combined network strength was computed for the test set, and the model was applied to generate ADOS predictions for these unseen participants.

Model performance was assessed^68^ by comparing the similarity between predicted and observed ADOS scores using both Spearman’s correlation (to avoid distribution assumptions)^134^. Note that performance was assessed on the entire sample after cross-validation. That is, in a given iteration, each of the ten folds resulted in a set of predictions for ADOS. After each participant had a predicted score from being in the test set, Spearman’s correlation was used to assess the relationships between predicted and observed scores across the entire sample. We performed 500 iterations of a given CPM analysis and selected the median-performing model; we report this in the main text when discussing model performance. To calculate significance, we randomly shuffled participant labels and attempted to predict ADOS scores. We repeated this 500 times and calculated the number of times a permuted predictive accuracy was greater than the median of the unpermuted predictions to achieve a non-parametric *P*-value:

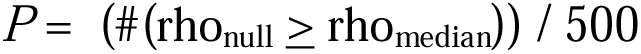

where #(rho_null_ ≥ rho_median_) indicates the number of permuted predictions numerically greater than or equal to the median of the unpermuted predictions^68^. We used the Benjamini–Hochberg method^135^ to correct for multiple comparisons, correcting for ten tests in the Yale youth sample (two for gradCPT, four for SSA, two for resting-state, one for gradCPT average, and one for resting-state average), three tests in the adult attention sample, and four tests in ABIDE.

We note the same CPM approach was used in the HBN sample, except 5-fold cross-validation was used, in line with the larger sample size^134^. All other parameters were the same as above. We corrected for three tests in the HBN sample when correcting for multiple comparisons.

### Testing generalizability of the ADOS network

To determine if the ADOS networks from the Yale youth sample generalized to external datasets (the adult attention sample, ABIDE, and HBN), we defined a consensus positive-association network and a consensus negative-association network as edges that appear in at least 6/10 folds in 300/500 iterations of CPM. This process resulted in 1,001 edges in the positive-association network and 1,013 edges in the negative-association network; hereafter, we refer to the collection of edges in the positive and negative networks as the ‘ADOS consensus network.’

We note the size of the ADOS consensus network is consistent with other CPM networks that have generalized^36,136,137^. To ensure generalizability results were robust, we tested summary networks of varying sizes (from liberal cases where an edge appeared in at least 1/10 folds and 50/500 iterations, to more stringent thresholds where an edge must appear in 10/10 folds and 500/500 iterations, moving in intervals of 1 fold and 50 iterations for each summary network).

To determine if the network predicted autistic traits, we then used the combined network strength in the ADOS consensus network and computed model coefficients across the Yale youth sample^36,67,74^. Model coefficients and the network masks were subsequently applied to the ABIDE and HBN samples to predict SRS scores. Model performance was determined by comparing the similarity between predicted and observed behavioral scores using Spearman’s correlation. We used the same approach to determine if the network predicted *d*’ scores.

To further assess generalizability, we repeated testing if the ADOS network predicts *d*’ and SRS using a multiverse approach. A multiverse analysis assesses how results are affected by different analytical choices^73^. Specifically, we tested if the ADOS positive and negative networks generalized; we adjusted models for IQ, age, and sex; and, as mentioned above, we tested a range of consensus network sizes. We point out the goal of a multiverse approach is not to determine what pipeline results in the highest prediction performance. Instead, the point is to assess various analytical scenarios and determine how different modelling choices impact generalization. As such, we do not perform multiple comparisons correction when assessing these results.

For completeness’ sake, we include the additional multiverse analyses performed in the Yale youth sample in this section. In this dataset, we adjusted CPM models for sex, age, and IQ; we also used CPM to predict social affect and restricted and repetitive behavior scores. Additionally, we assessed how altering the feature selection threshold impacted CPM.

### Testing generalizability of the SRS network from HBN

After determining that *Despicable Me* was the highest performing condition for SRS prediction, we reperformed CPM using the full 10 minutes of data from the functional scans (recall that comparing how scan condition affected CPM performance forced us to truncate all scans down to four minutes of data). After performing 500 iterations of CPM predicting SRS scores in HBN as before (median Spearman’s rho = 0.1398; *P* = 0.006), we generated a consensus network similar to above. Specifically, we required an edge to appear in at least 3/5 folds in 300/500 iterations of CPM. This process resulted in 940 edges in the positive-association network and 893 edges in the negative-association network. To ensure generalizability results were robust, we again tested summary networks of varying sizes (from liberal cases where an edge appeared in at least 1/5 folds and 50/500 iterations, to more stringent thresholds where an edge must appear in 5/5 folds and 450/500 iterations, moving in intervals of 1 fold and 50 iterations for each summary network. The resulting summary networks were then applied to the Yale youth sample to determine if the model generalized to predict ADOS scores using the gradCPT average data.

## Data availability

The functional parcellation is available here: (https://www.nitrc.org/frs/?group_id=51). ABIDE data are available here: (https://fcon_1000.projects.nitrc.org/indi/abide/). HBN data are available here: (https://fcon_1000.projects.nitrc.org/indi/cmi_healthy_brain_network/). To protect the sensitive nature of data in the adult attention and Yale youth sample, please reach out to the corresponding authors for questions about, or access to, these samples.

## Code availability

Preprocessing was carried out using freely available software: (https://medicine.yale.edu/bioimaging/suite/; this leads to the main page of BioImage Suite Web 1.2.0 (current build= 1.2.0, 2020/08/25, 11:43:02). Template scripts that were used for preprocessing of the functional data, as well as CPM code, are available here: (https://github.com/clhorien/tasks_versus_rest_in_autism_prediction/tree/main). CPM analyses were conducted in MATLAB (R2021b).

## Acknowledgements

This work was supported by the National Institutes of Health (P50MH115716, T32GM007205 to C.H. and A.S.G., R25MH119043 to C.H., and TR001864 to A.S.G.). The authors thank Hedwig Sarofin and Cheryl McMurray for assistance during the MRI sessions and Jitendra Bhawnani for assistance with hardware and software. The authors also thank Scuddy Fontenelle IV, Emma Brennan-Wydra, Chitra Banarjee, Rachel Foster, Veda Donthireddy, Kohrissa Joseph, Nicole Powell, Chaela Nutor, Diogo Fortes, and Maureen Butler for assistance with data collection and collation.

## Author Contributions Statement

CH conceptualized the study, with guidance from DS and RTC. CH performed the analyses with support from FM, ASG, XS, DO’C, EMRL, ESF, and RTC. CH, KC, and RTC designed the Yale youth study with support from KP, AV, JCM, FRV, DS; CH collected and processed the data for this study. MDR and MC designed the adult attention study; MDR collected and processed the data for this study. EMRL processed the data for the ABIDE study. BDA and LT processed the data for the HBN study. ESF and TS provided guidance on result interpretation. CH wrote the manuscript, with contributions from FM, ASG, EMRL, TS, and RTC, and comments from all authors.

