## Supplemental Materials for "Optimizing functional connectivity scanning conditions for predicting autistic traits"

**Supplemental Table of Contents**

Supplemental Methods……………………………………………………………………...……..2

Supplemental Table 1: Demographic comparisons between autism and non-autism participants in the Yale youth sample ……………………………………………...….….5

Supplemental Results………………………………………………………………………….....14

Supplemental Figure 1: Additional panels related to study design and control analyses.………………………………………………………………………………….14

Supplemental Table 2: CPM prediction performance when altering ADOS subscale or processing pipelines. ….………………………………………………………………....17

Supplemental Figure 2: Histograms showing the distributions of ADOS, SRS scores, and gradCPT scores in the Yale youth sample………………………………………...……..18

Supplemental Table 3: CPM prediction performance of different summary scores of autistic features. …………………………………………………………...………...….19

Supplemental Table 4: CPM prediction when altering processing pipelines and when performing case-control classification. ………………………………………………….20

Supplemental Table 5: Testing different ADOS network sizes and determining the relationship between predicted ADOS score and *d*’ in the adult attention sample.………………………………………………………………………….…….….21

Supplemental Table 6: Assessing generalizability of the ADOS network in ABIDE using different network sizes and when partialling out different variables…………...22

Supplemental Table 7 Testing different ADOS network sizes and determining prediction accuracy in HBN for SRS scores……………………………………………………..….23

Supplemental Table 8: Testing different DM consensus network sizes and determining the relationship between predicted SRS score and ADOS in the Yale youth sample. ….24

Supplemental Discussion………………………………………………………………….......…25

Supplemental References…………………………………………………………………...……28

**Supplemental Methods**

Recruitment, exclusion criteria of the Yale youth sample, and day-of-scan procedure

The neurodiverse sample has been described previously^1^. All participants and families provided informed consent for publication of participant data. As appropriate, we have adhered to the Strengthening the Reporting of Observational Studies in Epidemiology (STROBE) guidelines for reporting results related to observational studies[^50^](https://www.nature.com/articles/s41598-020-78885-z#ref-CR50).

Participants were recruited from the surrounding community through the use of flyers and other promotional materials. Specifically, recruitment materials were placed throughout the city of New Haven and surrounding areas in well-trafficked spaces (supermarkets, places of worship, community centers). In addition, participants were recruited from clinics at the Yale School of Medicine and the Yale Child Study Center that serve patients from different communities and with varying degrees of insurance. Yale’s Institutional Review Board approved the protocols for recruitment. In addition, in an effort to allow longitudinal analyses, participants from previous studies of autism in toddlers were contacted^2,3^. In total, 15/63 participants in the current work had previously taken part in studies of toddlers with autism. Additional work conducting longitudinal analyses in these participants is ongoing.

Participants were screened over the phone for basic developmental history and MRI safety factors. Those with a history of prematurity, known genetic abnormalities, suspected of having or having been diagnosed with hearing or visual impairment, or a history of non-febrile seizure disorders. Also, we screened for those having metal implants or other devices that are not MR compatible, and children with braces were excluded to ensure imaging quality was unaffected.

Consistent with the dimensional approach used here and aligned with the Research Domain Criteria, we specifically wanted to capture a heterogenous sample that varied along multiple dimensions^4^. In the context of the current work, such an approach aims to more accurately capture the spectrum of functioning as it pertains to autism and helps to alleviate concerns associated with the use of a classic case-control approach^4-6^. Post-hoc analyses revealed that despite our focus on acquiring a heterogenous sample, those diagnosed with autism were well-matched with non-autistic participants on a variety of measures (Supplemental Table 1). Consistent with the 3:1 sex ratio of autism diagnoses amongst males:females^7,8^, the autism participants tended to be male and also tended to be slightly older than the non-autistic participants. To ensure these differences were not driving our findings, we adjusted for these variables when performing CPM and observed no impact on performance

(see Supplemental Table 3).

One hundred and two participants were scanned. We required all study participants to pass visual quality control (QC) after preprocessing (i.e. all skull-stripped data, linear registrations, and non-linear registrations were manually inspected), as well as to have data from all eight functional runs. We excluded participants with a lack of full brain coverage during functional scans (n = 7). Eight participants had an imaging artifact preventing proper registration. Fifteen participants concluded the study early. One participant was excluded due to ending the final rest run early. Participants were excluded due to a technical issue with the task laptop preventing proper recording of button presses (n = 7) and a participant (n = 1) was excluded due to pressing a combination of buttons during the gradCPT. This left 63 participants who passed visual QC and had all functional scans for all main analyses. Three of the participants with autism did not have information regarding secondary diagnoses; we thus used a sample size of 60 when comparing secondary diagnoses among those with autism versus those without. In addition, six of the 63 participants (1 participant with autism) did not have usable gradCPT *d*’ behavioral data due to a technical factor impairing button presses during gradCPT performance; we used a sample size of 57 participants when comparing gradCPT performance among those with autism versus those without.

Each participant was clinically characterized at Dr. Katarzyna Chawarska’s Yale Early

Social Cognition Lab. All imaging was completed at the Yale Magnetic Resonance Research Center (MRRC), directed by Dr. R. Todd Constable. Participants were clinically characterized prior to the MRI scanning session (depending on participant availability, this ranged from 1-2 weeks prior to the MRI session to 4-5 days prior to the session). Diagnostic classification of autism was based on clinical best estimate diagnosis by a team of clinical psychologists and any available reports of developmental and medical history, along with ADOS scores. Participants completed behavioral questionnaires and other clinical assessments approximately 4-5 days prior to the scanning session.

All clinicians involved in this study have undergone extensive reliability training, with special attention devoted to diagnostic procedures (especially the ADOS-2, used here). Monthly meetings were conducted to discuss potential issues related to clinical characterization and maintain consistency of data collection and research procedures. In addition, double-scoring and double-entry of protocols were performed, with immediate checks for discrepancies and resolution by the support staff. In addition, a clinical social worker was available to all families throughout their length of study participation to offer assistance or to provide access to resources in case needs arose.

Participants conducted a mock scan session approximately 4-5 days before the MRI scanning session. We have described the mock scan session previously^1^. Briefly, the session lasted about an hour and acclimated the participants to the scanning environment and comprised a prize system based on real-time estimates of head motion. The day of the scan, participants completed a brief mock scan refresher session, lasting about 20 minutes. The MRI scanning session also used a prize system to encourage participants to minimize head motion.

| Measure | Autism participants | Non-autism participants | *t*-statistic | *P*-value (corrected) |
| --- | --- | --- | --- | --- |
| gradCPT performance (mean, SD) | 2.59 (0.96) | 2.66 (0.86) | -0.27 | 0.79 |
| Age | 13.2 (3.12) | 11 (2.31) | 3.12 | 0.0028* |
| IQ | 103.5 (17.3) | 109.8 (13.8) | -1.56 | 0.1236 |
| Head motion | 0.0962 (0.0486) | 0.0991 (0.0415) | -0.24 | 0.8089 |
| Sex (males) | 15 | 19 | 4.05^ | 0.044 |

Supplemental Table 1. Demographic comparisons between autism and non-autism participants in the Yale youth sample. All *P*-values computed using a two-sample t-test (^except sex, which used a chi-square test, with Yates correction). Asterisk indicates significant after multiple comparisons correction using the Benjamini–Hochberg method^9^.

Acquisition parameters of the Yale youth sample

Participants were scanned on a 3T Siemens Prisma system. After a localizer, participants underwent the following scans: anatomical magnetization prepared rapid gradient echo (MPRAGE); T1 fast low angle shot (T1 FLASH); two runs of gradCPT; four movie runs; two runs of resting-state runs; and T2-weighted 3D fast spin echo image (T2 SPACE).

MPRAGE images were obtained with the following image parameters: 208 contiguous slices acquired in the sagittal plane, repetition time (TR) = 2400 ms, echo time (TE) = 1.22 ms, flip angle = 8°, slice thickness = 1 mm, in-plane resolution = 1 mm × 1 mm, matrix size = 256 × 256.

T1 FLASH images were obtained with the following sequence parameters: 75 contiguous slices acquired in the axial-oblique plane parallel to AC-PC line, TR = 440 ms, TE = 2.61 ms, flip angle = 70°, slice thickness = 2 mm, in-plane resolution = 0.9 mm × 0.9 mm, matrix size = 256 × 256.

T2 SPACE images were obtained with the following sequence parameters: 208 slices per slab acquired in the sagittal plane, TR = 3200 ms, TE = 316 ms, slice thickness = 1 mm, in-plane resolution = 1 mm x 1 mm, matrix size = 256 x 256 x 208.

Functional images were acquired using a multiband gradient echo-planar imaging (EPI) pulse sequence with the following image parameters: 75 contiguous slices acquired in the axial-oblique plane parallel to the AC-PC line, TR = 1000 ms, TE 30 ms, voxel size = 2.0 mm^3^, flip angle = 55 degrees, slice thickness = 2 mm, bandwidth = 1894 Hz/pixel, matrix size = 110 x 110, field of view = 220 mm, multiband factor = 5.

Acquisition parameters of the adult attention sample

Participants from a previously published study were scanned on a 3T Siemens Trio TIM system (note this is a different scanner than used to collect data in the Yale youth sample; also note the acquisition sequences were different than those used in the Yale youth sample). MPRAGE images were obtained with the following image parameters: TR = 2530 ms, TE = 3.32, flip angle = 7°, acquisition matrix = 256 × 256, in-plane resolution = 1.0 mm^2^, slice thickness = 1.0 mm, 176 sagittal slices.

Functional images were acquired using a multiband gradient echo-planar imaging (EPI) pulse sequence with the following image parameters: (TR) = 1,000 ms, echo time (TE) = 30 ms, flip angle = 62°, acquisition matrix = 84 × 84, in-plane resolution = 2.5 mm^2^, 51 axial-oblique slices parallel to the ac-pc line, slice thickness = 2.5, multiband 3, acceleration factor = 2, total whole-brain volumes obtained = 824. A 2D T1-weighted image with the same slice prescription as the EPI images was also obtained for registration.

Imaging details and exclusion criteria of the ABIDE sample

We used the subjects described in Lake et al.^10^. From this pool of subjects (n = 352 with low-motion data, < 0.10 mm mean frame-to-frame displacement; FFD), we additionally excluded subjects who had incomplete scan coverage (i.e., many subjects did not have adequate coverage of the cerebellum, brainstem, and dorsal portions of cortex, thus resulting in missing significant numbers of nodes from the functional parcellation used here). This resulted in the exclusion of 123 subjects with low-motion, full coverage brain data, leaving a final sample of n = 229. Of the original pool of 58 subjects with ADOS data from Lake et al.^10^, we excluded 25 due to a lack of full-brain coverage, resulting in a final sample size of 33 low-motion, full-brain coverage subjects. Hence, we chose to analyze the SRS pool of subjects given the larger number of subjects compared to ADOS. See Lake et al.^10^ for imaging parameters of the ABIDE subjects.

Imaging details and exclusion criteria of the HBN sample

This was a multi-site study; scanning took place using a mobile MRI scanner in Staten Island, as well as the CitiGroup Cornell Brain Imaging Center, the Rutgers University Brain Imaging Center, and the CUNY Advanced Science Research Center. Structural images included MPRAGE images, as well as T2 fluid attenuated inversion recovery (FLAIR) images. Functional images were obtained while participants completed two rest fMRI runs as well as *Despicable Me* and *The Present* movie-watching scan sessions. We used the exclusion criteria described in Adkinson et al^11^, with a mean FFD threshold of < 0.2 mm imposed across scans. Of the 643 individuals that remained, we used the Schedule for Affective Disorders and Schizophrenia—Children’s version (KSADS)^12^ to determine diagnosis. Specifically, we used parent and clinician assessment for autism (n = 107) and clinician assessment for ADHD diagnoses (n = 301). See Alexander et al.^13^ for imaging parameters of the HBN sample. As mentioned in the main text, we used data from 643 participants in the current manuscript when assessing if the ADOS model generalized in HBN. When conducting CPM analyses that required the truncation of time courses, we excluded one subject (NDARRM725BRV), for a final sample size of 642 (due to missing 210 frames in resting-state run 1).

Limiting the impact of head-motion

Participant head motion while being scanned is known to affect functional connectivity^14-16^ and brain–behavior relationships^17^. This fact must be considered in any study attempting to link brain connections to clinical phenotypes, including the present work. To limit the impact of motion, we undertook numerous steps in the current work. First, all participants in the Yale youth sample participated in a mock scan protocol approximately a week before the MRI session. In other work, we have demonstrated in the Yale youth sample that the mock scan protocol results in a statistically significant reduction of in-scanner head motion^1^. In addition, we have demonstrated in the same sample that robust connectome-based markers of phenotypes can be generated and that such markers are not driven by head motion^18^.

In the present work, the mock scan protocol led to approximately 90% of the sample having scans below the 0.25 mm mean FFD threshold (see Supplemental Figure 1G below), a motion threshold in line with those used for determining high- versus low-motion data in youth and/or those with a psychiatric condition^19-21^. We also point out that the number of participants with scans below 0.25 mm mean FFD is much lower compared to other samples of youth^22^. In addition, the data from the adult attention sample, the ABIDE data, and the HBN data are low motion. Specifically, all subjects from the adult attention sample had a mean FFD below 0.087 mm, all subjects used from ABIDE had a mean FFD below 0.10 mm, and all subjects from HBN had a mean FFD below 0.20 mm.

As in previous brain-behavior studies using gradCPT^23,24^, no censoring of the functional data was performed. Thus, this allowed us to avoid the confound of having different numbers of timepoints and button presses across participants (a concern when using the present gradCPT fMRI data, given that the calculation of *d’* scores is via second-to-second button presses). Furthermore, as in other CPM papers^10,18,25,26^, additional steps were taken during CPM to limit the potential of head motion causing spurious brain-behavior relationships. In particular, we incorporated the participants’ mean FFD during a given functional scan into the CPM model and observed that motion did not seem to affect predictions (e.g., using gradCPT as an example, the model still successfully predicted ADOS in the Yale youth sample when controlling for motion; gradCPT average: Spearman’s rho = 0.445, *P*-value = 0.002, corrected). In addition, when testing if the ADOS network model generalized in the adult attention sample, we again adjusted the model for mean FFD and determined that results were not being driven by head motion (e.g., performance was unaffected; Spearman’s rho = -0.56, *P*-value = 0.0043). Similarly, when we adjusted the ADOS models for head motion in ABIDE and HBN, predictions of SRS scales remained high (Supplemental Table 6 for ABIDE prediction performance; prediction performance in HBN when controlling for head motion: Spearman’s rho = 0.1002, *P* = 0.0111).

Moreover, the fact that the ADOS model from the Yale youth sample generalized in the adult attention sample when both have different stimuli timing parameters increases confidence that the model’s success in the adult attention sample is not due to artifactual head movements linked to stimuli presentation^18^. That is, gradCPT stimuli were shown every 1000 ms in the Yale youth sample and every 800 ms in the adult attention sample. If there was overfitting in the Yale youth sample due to motion artifact, this would hinder prediction of *d*’ in the adult attention sample. We also point out that the prediction of SRS in ABIDE and HBN further supports that motion-induced artifacts are not driving predictions in the current work. Hence, the fact that the model predicts out of sample in three independent datasets suggests that head motion is not driving the brain-behavior model observed here.

To summarize, head motion is known to be a concern when determining brain–behavior relationships. Nevertheless, the present data suggest that motion is not driving the conclusions of the current paper.

Counterbalancing the SSA clips and study design considerations of the Yale youth sample

We note here a few points of clarification regarding counterbalancing of the SSA clips (Supplemental Figure 1A) and general study design. Each eye contact and speech condition required counterbalanced SSA data. To achieve this, we used four separate conditions (referred to as A, B, C, and D for brevity). Each two-minute clip was shown twice across four scans. Furthermore, to allow adequate sample size in case of participant attrition, we showed each of the four conditions once during the first 2/4 SSA functional runs; we then showed each of the four conditions once again during the final 2/4 SSA functional runs. Specifically, for a given participant, conditions A, B, C, and D were always shown during the first two functional runs; A, B, C, and D were then shown in the final two functional runs. The clip presentation was then counterbalanced across participants. This strategy was chosen so that even if a participant ended the study after obtaining only 2/4 SSA functional runs, they would still have all 4 SSA conditions (albeit with only two minutes of data).

In addition, it is well known that scanning youth^27^, especially youth with a neurodevelopmental condition^28,29^, is difficult due to both participant head motion and attrition^30^. As such, most studies collecting functional MRI data in youth with autism tend to only collect 1-2 functional scans per participant. Short MRI protocols like these have the advantage of allowing data collection from subjects who are challenging to scan. However, due to the low amount of data per participant, these studies tend to have low-reliability functional connectivity data^31,32^. To increase reliability, more scanning data are needed per subject.

Nevertheless, the realities of collecting a well-powered study with adequate data per subject, along with a sufficient number of subjects in the entire study, force investigators to make trade-offs in study design^33^. Based on pilot data in the Yale youth sample, in which we found subjects had high motion data^1^ and tended to abort the study early, we collected data in the following order: gradCPT scans, then SSA tasks, then resting-state scans. We chose this order as we wanted to maximize the number of scans collected across the highest number of subjects, ensuring that our sample was both ‘broad’ and ‘deep’^34^. For instance, we reasoned that if all participants chose to end the study early during SSA functional run three, then there would be data from two gradCPT scans and at least conditions A-D of the SSA task. Note that this is a real concern in neurodevelopmental studies. In the current study, as an example, we required a participant to have data from all eight functional scans, across all three scanning conditions. Thus, there were 16/102 subjects who chose to leave the scanner early and were excluded.

As we note in the main text, it is possible the SSA task and resting-state conditions may have underperformed in terms of prediction performance as they came later in the study, and participants may have been fatigued. However, as others have shown^35^, increased prediction performance of task versus rest data does not seem to be related to arousal, but rather due to cognitive differences elicited by the task^35^. Future studies could more clearly delineate the role of arousal in brain-behavior relationships in autism.

There are strengths and limitations to having an established functional run order across subjects. We simply point out that all of these design considerations should be kept in mind when interpreting the results of the current work and should also be considered when investigators are designing their own MRI protocols comparing task conditions. We direct the curious reader to our previous work, where we detail other practical components associated with collecting high-quality low-motion data in this population.^1^

Connectome-based predictive modelling

CPM (Supplemental Figure 1B) was used to generate brain-behavior models. Specifically, using 10-fold cross-validation, connectivity matrices from a given scan condition and ADOS scores were split into an independent training set including subjects from 9 folds and a test set including the left-out fold. Linear regression was used to relate edge strength to ADOS score in the training set. Edges most strongly associated with ADOS scores were selected (feature selection threshold of *P* = 0.05) for both a ‘positive network’ (in which increased connectivity was associated with higher ADOS scores) and a ‘negative network (in which decreased connectivity was associated with lower ADOS scores). We used partial correlation to control for mean participant head motion at the feature selection step^18,26,36^. Mean network strength was computed in both the positive and negative networks, and the difference between these network strengths was computed (‘combined network strength’), as in previous work ^37^: A linear model was then calculated relating combined network strength to ADOS scores in the training set. In the last step, combined network strength was computed for the test set, and the model was applied to generate ADOS predictions for these unseen participants.

Model performance was assessed as reported previously ^26^ by comparing the similarity between predicted and observed ADOS scores using both Spearman’s correlation (to avoid distribution assumptions)^38^. We performed 500 iterations of a given CPM analysis and selected the median-performing model; we report this in the main text when discussing model performance. To calculate significance, we randomly shuffled participant labels and attempted to predict ADOS scores. We repeated this 500 times and calculated the number of times a permuted predictive accuracy was greater than the median of the unpermuted predictions to achieve a non-parametric *P*-value:

*P* = (#(rho_null_ ≥ rho_median_)) / 500

where #(rho_null_ ≥ rho_median_) indicates the number of permuted predictions numerically greater than or equal to the median of the unpermuted predictions^26^. We used the Benjamini–Hochberg method^9^ to correct for multiple comparisons, correcting for ten tests in the Yale youth sample, three tests in the adult attention sample, and four tests in ABIDE.

**Supplemental Results**

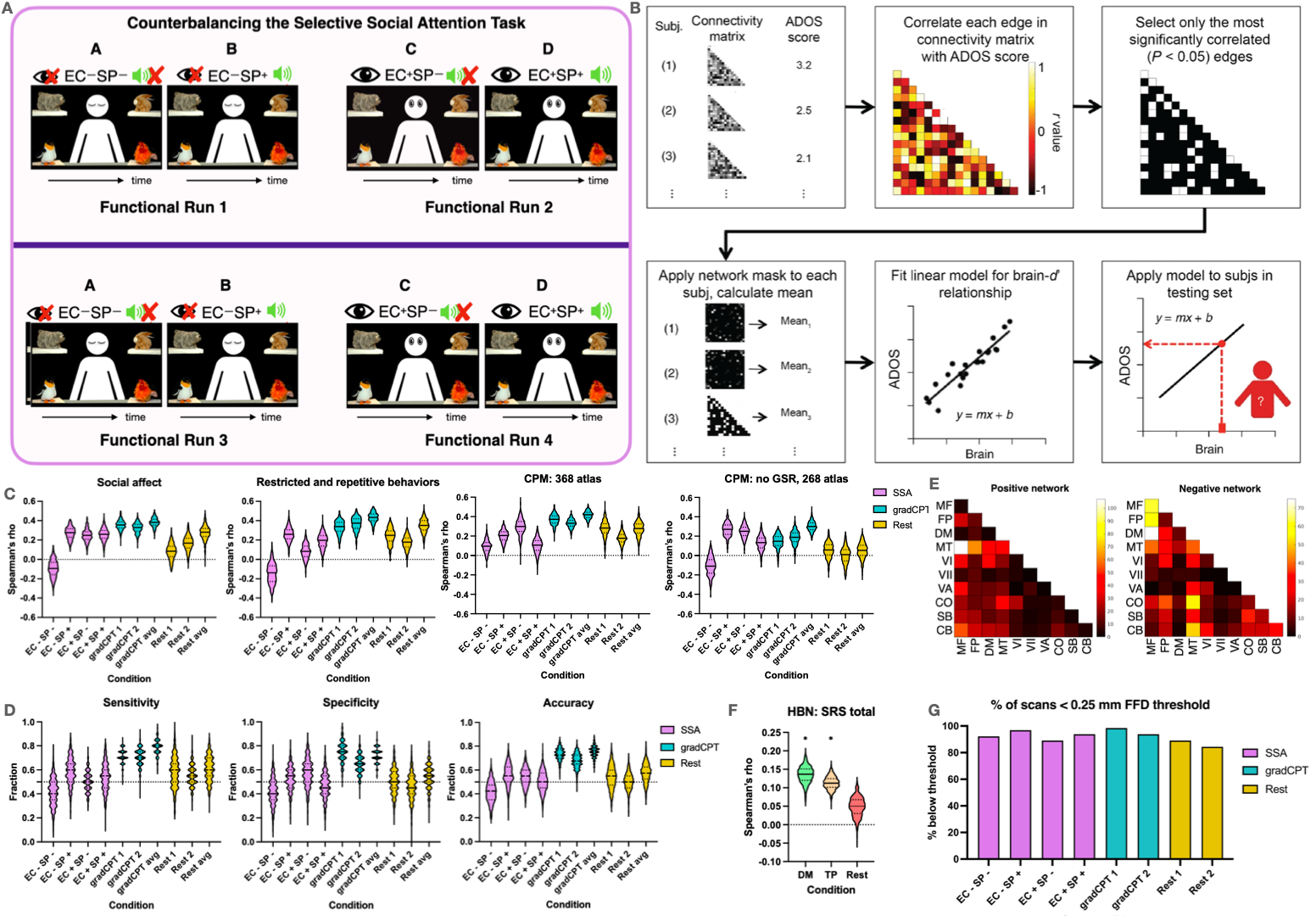

Supplemental Figure 1. Additional panels related to study design and control analyses.

A) Counterbalancing the SSA task. We showed participants images from four separate conditions, stratified according to speech and eye contact, which we abbreviate here for simplicity. Condition A = no eye contact, no speech (EC-SP-). Condition B = no eye contact, with speech (EC-SP+). Condition C = eye contact, no speech (EC+SP-). Condition D = eye contact, with speech (EC+SP+). In a single functional run, participants were shown two clips, separated by a fifteen-second fixation cross. The order was such that participants were shown clips A-D spread out across the first two functional runs, in a counterbalanced order across subjects (top part of figure, above the dark purple line in the middle of the figure). Participants were then shown clips A-D again during functional runs 3 and 4, in a counterbalanced order across subjects (bottom part of figure, below the dark purple line). As described above, this served to maximize data collection across the four separate conditions in case of participant attrition. Please refer to the Methods for a more thorough description of each individual SSA condition.

B) A schematic detailing the step-by-step process of CPM. Figure adapted with permission from Shen et al. (2017). Note we have included text above describing CPM that is also in the Methods; we include it here to aid interpretation of the Figure.

C) CPM prediction performance when altering ADOS subscale or processing pipelines. For all plots, the scan condition is shown on the x-axis; on the y-axis, Spearman’s rho is shown for the correlation of the predicted ADOS scores with actual ADOS scores. For each condition, the median of the 500 iterations is shown as a solid black line in the violin plot; quartiles, as dotted lines. The Selective Social Attention task conditions are shown in purple, gradCPT is shown in turquoise, and resting-state data in yellow. Note the same data are included below in Supplemental Tables 2 and 4; please see these tables for exact *P*-values. Statistical significance was obtained using permutation testing as described in the Methods. Note that one-side significance testing was used, as we are interested in statistically significant positive predictions only. The Benjamini–Hochberg method^9^ was used to control for multiple comparisons. Results obtained from n = 63 participants.

D) Case-control classification of autism diagnosis. CPM was modified to predict whether a participant had autism or not. Sensitivity (the number of those correctly classified with autism / total number of autism participants) is shown to the left. Specificity (the number of those correctly classified as non-autism / total number of non-autism participants) is shown in the middle. Accuracy (the number of correct classifications / all classifications) is shown to the right. The scan condition is shown on the x-axis; on the y-axis, the fraction is shown, with 0 indicating no correct classifications and 1 indicating all participants were correctly classified. A dotted line is shown at chance level, in this case at 0.5. For each condition, the median of the 500 iterations is shown as a solid black line in the violin plot; quartiles, as dotted lines. The Selective Social Attention task conditions are shown in purple, gradCPT in turquoise, and resting-state data in yellow. Note that to ensure balanced cases and controls, we subsampled the non-autistic participants in each iteration such that there were 20 cases of autism and 20 cases of controls. Note the same data are included below in Supplemental Table 4. Please see the Tables for exact *P*-values. Statistical significance was obtained using permutation testing as described in the Methods. Note that one-side significance testing was used, as we are interested in statistically significant positive predictions only. The Benjamini–Hochberg method^9^ was used to control for multiple comparisons.

E) Neuroanatomy of predictive edges in the ADOS model. Note these are the same data as shown in Figure 6C-D of the main text, except the raw number of edges per network pair is shown. Left matrix): matrix of the positive-association network. Right matrix): matrix of the negative-association network. Network labels: MF, medial frontal; FP, frontoparietal; DM, default mode; MT, motor; VI, visual I; VII, visual II; VA, visual association; CO, cingulo-opercular; SB, subcortical; CB, cerebellum

F) Testing for convergence of task versus rest CPM findings in HBN. CPM prediction performance across different scanning conditions for SRS scores in the Healthy Brain Network. The scan condition is shown on the x-axis; on the y-axis, Spearman’s rho is shown for the correlation of predicted and actual SRS scores. For each condition, the median of the 500 iterations is shown as a solid black line in the violin plot; quartiles, as dotted lines. Asterisk (*) indicates statistical significance after correcting for multiple comparisons. Statistical significance was obtained using permutation testing as described in the Methods. Note that one-side significance testing was used, as we are interested in statistically significant positive predictions only. The Benjamini–Hochberg method^9^ was used to control for multiple comparsions. Exact P-values from statistically significant conditions, median performing model from Despicable Me: Spearman’s rho = 0.14, *P*-value < 0.001, corrected; median performing model from The Present: Spearman’s rho = 0.11, *P*-value < 0.0001, corrected. Results obtained from n = 643 participants.

G) Quantifying head motion in the Yale youth sample. The percentage of scans below a 0.25 mm mean FFD threshold. The SSA task conditions are shown in purple, gradCPT is shown in turquoise, and resting-state data in yellow. Results obtained from n = 63 participants.

Abbreviations: ADOS, autism diagnostic observation schedule; Avg, average; DM, Despicable Me; EC-SP-, no eye contact, no speech; EC-SP+, no eye contact, with speech; EC+SP-, eye contact, with no speech; EC+SP+, eye contact, with speech; HBN, Healthy Brain Network; mm, millimeters; SSA, selective social attention task; SRS, social responsiveness scale; TP = The Present.

CPM prediction performance when altering ADOS subscale or processing pipelines

|  | CPM prediction performance of ADOS scales | | | | | |  | CPM prediction performance of ADOS total severity score when partialling out different variables | | | | | |
| --- | --- | --- | --- | --- | --- | --- | --- | --- | --- | --- | --- | --- | --- |
|  | **Social affect** | | **RRB** | | **Total ADOS** | |  | **Sex** | | **Age** | | **IQ** | |
|  | Rho | *P*-val | Rho | *P*-val | Rho | *P*-val |  | Rho | *P*-value | Rho | *P*-value | Rho | *P*-value |
| **SSA: EC-SP-** | -0.093 | 0.814 | -0.139 | 0.888 | -0.106 | 0.822 |  | -0.089 | 0.788 | -0.067 | 0.704 | -0.174 | 0.926 |
| **SSA: EC+SP-** | 0.276 | 0.046 | 0.258 | 0.074 | 0.251 | 0.052 |  | 0.114 | 0.208 | 0.119 | 0.224 | 0.219 | 0.116 |
| **SSA: EC-SP+** | 0.249 | 0.052 | 0.083 | 0.298 | 0.266 | 0.034 |  | 0.146 | 0.186 | 0.191 | 0.072 | 0.289 | 0.016 |
| **SSA: EC+SP+** | 0.259 | 0.038 | 0.198 | 0.11 | 0.128 | 0.148 |  | 0.03 | 0.402 | 0.075 | 0.282 | 0.053 | 0.36 |
| **gradCPT 1** | 0.356 | 0.016 | 0.339 | 0.022 | 0.441 | 0.004* |  | 0.348 | 0.016 | 0.292 | 0.036 | 0.427 | 0.006 |
| **gradCPT 2** | 0.33 | 0.036 | 0.375 | 0.006 | 0.35 | 0.018 |  | 0.307 | 0.068 | 0.291 | 0.022 | 0.338 | 0.024 |
| **gradCPT avg** | 0.38 | 0.014 | 0.438 | 0.008 | 0.445 | 0.002* |  | 0.373 | 0.018 | 0.319 | 0.03 | 0.452 | 0.0001 |
| **Rest 1** | 0.084 | 0.27 | 0.249 | 0.046 | 0.093 | 0.23 |  | 0.002 | 0.522 | 0.175 | 0.136 | 0.069 | 0.262 |
| **Rest 2** | 0.169 | 0.138 | 0.178 | 0.096 | 0.18 | 0.124 |  | 0.106 | 0.206 | 0.172 | 0.116 | 0.163 | 0.124 |
| **Rest avg** | 0.28 | 0.04 | 0.35 | 0.014 | 0.296 | 0.038 |  | 0.131 | 0.19 | 0.25 | 0.058 | 0.231 | 0.058 |

Supplemental Table 2. CPM prediction performance when altering ADOS subscale or processing pipelines. Left portion of table: CPM prediction performance of ADOS scales. Shown is the median Spearman’s rho of the correlation between actual and predicted ADOS scores after completing 500 iterations. Note the same data for social affect and RRB are included in Supplemental Figure 1C. Right portion of the table: CPM prediction performance of total ADOS severity score when partialling out different variables (indicated by the different columns). Shown is the median Spearman’s rho of the correlation between actual and predicted ADOS scores after completing 500 iterations. The *P*-value is shown as determined from permutation testing (Methods). Asterisk (*) indicates statistical significance. Note that one-side significance testing was used, as we are interested in statistically significant positive predictions only. The Benjamini–Hochberg method^9^ was used to control for multiple comparisons. ADOS, autism diagnostic observation schedule; Avg, average; EC-SP-, no eye contact, no speech; EC-SP+, no eye contact, with speech; EC+SP-, eye contact, with no speech; EC+SP+, eye contact, with speech; *P*-val, *P*-value; RRB, restricted and repetitive behaviors; SSA, selective social attention.

Distributions of different phenotypes of interest in the Yale youth sample

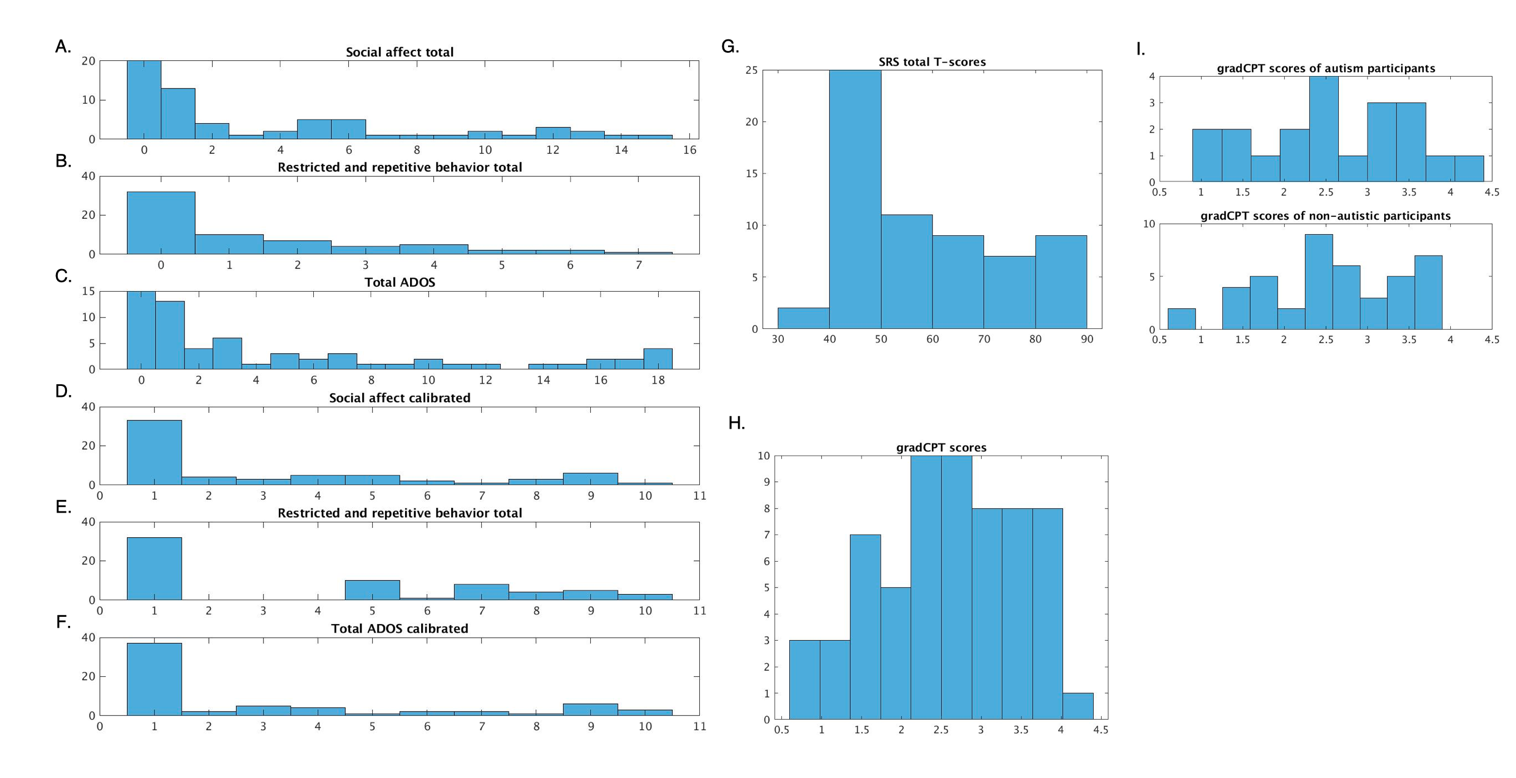

Supplemental Figure 2. Histograms showing the distributions of ADOS, SRS scores, and gradCPT scores in the Yale youth sample. The top three rows (A-C) of the leftmost column comprise data from the raw subscales, whereas the bottom three rows (D-F) of the leftmost column comprise data from the calibrated scales. G. SRS total T-scores. H. The gradCPT scores (*d*’) from all participants in the sample. I. The gradCPT scores from participants split by diagnosis. Results obtained from n = 63 participants. ADOS, autism diagnostic observation schedule; SRS, social responsiveness scale.

CPM prediction of additional phenotypes in the Yale youth sample

| **SRS total T-scores** | | **ADOS total score (uncalibrated)** | | **ADOS total score (calibrated), balancing autistic and non-autistic participants** | | **gradCPT performance** | |
| --- | --- | --- | --- | --- | --- | --- | --- |
| Rho | *P*-value | Rho | *P*-value | Rho | *P*-value | Rho | *P*-value |
| 0.39 | 0.008 | 0.4 | 0.004 | 0.41 | 0.006 | 0.26 | 0.042 |

Supplemental Table 3. CPM prediction performance of different summary scores of autistic features. Shown is the median Spearman’s rho of the correlation between the actual and predicted phenotype of interest after completing 500 iterations using the gradCPT average data in the Yale youth sample. CPM parameters were kept identical to those used in the main text, including the partialling of participant head motion. For the column reading **“**ADOS total score, balancing autistic and non-autistic participants,” we performed CPM after subsampling the non-autistic participants in each iteration such that there were 20 cases of autism and 20 cases of controls. The *P*-values were determined from permutation testing (Methods) and are shown for context. Note that due to the post-hoc nature of this analysis, we are not interested in statistical significance of the new CPM pipelines per se. Rather, we are interested in the overall pattern of results and determining if results generally converge across the different pipelines, consistent with the multiverse approach used elsewhere in the paper^39^. ADOS, autism diagnostic observation schedule; SRS, social responsiveness scale.

CPM prediction when altering processing pipelines and when performing case-control classification

| Alternative CPM modelling context |  | Model performance | |  | Model performance |  |  | Sens |  | Spec |  | Acc | |
| --- | --- | --- | --- | --- | --- | --- | --- | --- | --- | --- | --- | --- | --- |
|  | Model | Spearman’s rho | *P*-value |  | Spearman’s rho | *P*-value |  | Median | *P*-value | Median | *P*-value | Median | *P*-value |
| 368 atlas | EC- SP - | 0.095 | 0.678 | No GSR | -0.11 | 0.794 | Case-control classification | 0.4 | 0.454 | 0.4 | 0.716 | 0.425 | 0.586 |
|  | EC- SP + | 0.205 | 0.184 |  | 0.27 | 0.136 |  | 0.6 | 0.434 | 0.55 | 0.7 | 0.55 | 0.546 |
|  | EC+ SP - | 0.3 | 0.001 |  | 0.251 | 0.01 |  | 0.5 | 0.416 | 0.6 | 0.758 | 0.55 | 0.558 |
|  | EC+ SP + | 0.107 | 0.042 |  | 0.133 | 0.678 |  | 0.55 | 0.432 | 0.45 | 0.722 | 0.5 | 0.546 |
|  | gradCPT 1 | 0.372 | 0.012 |  | 0.148 | 0.214 |  | 0.7 | 0.128 | 0.75 | 0.042 | 0.725 | 0.028 |
|  | gradCPT 2 | 0.332 | 0.066 |  | 0.189 | 0.108 |  | 0.7 | 0.112 | 0.65 | 0.17 | 0.675 | 0.052 |
|  | gradCPT avg | 0.42 | 0.012 |  | 0.3 | 0.046 |  | 0.8 | 0.03 | 0.7 | 0.094 | 0.75 | 0.018 |
|  | Rest 1 | 0.283 | 0.02 |  | 0.058 | 0.394 |  | 0.6 | 0.252 | 0.5 | 0.61 | 0.55 | 0.32 |
|  | Rest 2 | 0.177 | 0.096 |  | 0.009 | 0.43 |  | 0.55 | 0.398 | 0.45 | 0.716 | 0.5 | 0.56 |
|  | Rest avg | 0.278 | 0.018 |  | 0.053 | 0.386 |  | 0.6 | 0.258 | 0.55 | 0.376 | 0.575 | 0.21 |

Supplemental Table 4. CPM prediction when altering processing pipelines and when performing case-control classification. Leftmost columns: results obtained using a 368-node atlas. Middle columns: results obtained using a 268-node atlas with no GSR. Rightmost columns: results obtained from case-control classification. All columns: The median-performing model after 500 iterations is shown. *P*-values for the alternative CPM modelling pipelines were obtained from permutation testing and are shown for context. Note that due to the post-hoc nature of this analysis, we are not interested in statistical significance of the new CPM pipelines per se. Rather, we are interested in the overall pattern of results and determining if results generally converge across the different pipelines, consistent with the multiverse approach used elsewhere in the paper^39^. Note: these same data appear in Supplemental Figures 1C and 1D. Acc, accuracy; ADOS, autism diagnostic observation schedule; Avg, average; EC-SP-, no eye contact, no speech; EC-SP+, no eye contact, with speech; EC+SP-, eye contact, with no speech; EC+SP+, eye contact, with speech; *P*-val, *P*-value; RRB, restricted and repetitive behaviors; SSA, selective social attention; Sens, sensitivity; Spec, specificity.

Testing different ADOS network sizes in the adult attention sample

|  | | | | **Results using positive network** | | **Results using negative network** | | **Results using combined network** | |
| --- | --- | --- | --- | --- | --- | --- | --- | --- | --- |
| **Parameters tested (folds, iterations)** | **Positive network size** | **Negative network size** | **Combined network size** | **Rho** | ***P*-value** | **Rho** | ***P*-value** | **Rho** | ***P*-value** |
| 1, 50 | 5966 | 5812 | 11,778 | -0.54 | 0.0061 | -0.56 | 0.004 | -0.54 | 0.0059 |
| 2, 100 | 3119 | 3051 | 6,170 | -0.56 | 0.0039 | -0.56 | 0.0042 | -0.57 | 0.0035 |
| 3, 150 | 2181 | 2176 | 4,357 | -0.57 | 0.0033 | -0.49 | 0.0132 | -0.54 | 0.0057 |
| 4, 200 | 1670 | 1651 | 3,321 | -0.58 | 0.0027 | -0.45 | 0.0245 | -0.56 | 0.0042 |
| 5, 250 | 1313 | 1261 | 2,574 | -0.59 | 0.0023 | -0.46 | 0.0217 | -0.55 | 0.0047 |
| 6, 300 | 1001 | 1013 | 2,014 | -0.59 | 0.0021 | -0.46 | 0.0230 | -0.56 | 0.0049 |
| 7, 350 | 773 | 763 | 1,536 | -0.59 | 0.0025 | -0.45 | 0.0268 | -0.55 | 0.0048 |
| 8, 400 | 532 | 533 | 1,065 | -0.53 | 0.0073 | -0.41 | 0.0441 | -0.50 | 0.0111 |
| 9, 450 | 283 | 277 | 560 | -0.60 | 0.002 | -0.21 | 0.3195 | -0.42 | 0.0365 |
| 10, 500 | 26 | 30 | 56 | -0.43 | 0.0344 | -0.26 | 0.2170 | -0.30 | 0.1397 |

Supplemental Table 5. Testing different ADOS network sizes and determining the relationship between predicted ADOS score and *d*’ in the adult attention sample. To generate each summary network, we required that an edge appear in *n*/10 k-folds and *j*/500 CPM iterations (leftmost column). Specifically, the variables *n* and *j* were initially set at 1 and 50, respectively. The variables were altered in increments of 1 and 50, respectively. The rows of the table are arranged such that the top row is the most liberal network summarization approach (i.e., an edge had to appear in only 1/10 folds and 50/500 iterations); the bottom of the table is the most stringent case (i.e., an edge must appear in 10/10 folds and 500/500 iterations). Rho = Spearman’s rho of the correlation between predicted ADOS scores and *d*’ scores. *P*-values (associated with the corresponding Spearman’s rho) for the alternative network sizes are shown for context. Note that due to the post-hoc nature of this analysis, we are not interested in statistical significance of the new CPM pipelines per se. Rather, we are interested in the overall pattern of results and determining if results generally converge across the different pipelines, consistent with the multiverse approach used elsewhere in the paper^39^.

Assessing generalizability of the ADOS network in ABIDE using different network sizes and when partialling out different variables

|  |  | **SRS total scores** | | **SRS communication scores** | | **SRS motivation scores** | | **SRS mannerisms scores** | |
| --- | --- | --- | --- | --- | --- | --- | --- | --- | --- |
|  | **Parameters tested** | **Rho** | ***P*-value** | **Rho** | ***P*-value** | **Rho** | ***P*-value** | **Rho** | ***P*-value** |
|  | 1, 50 | 0.19234 | 0.00348 | 0.16200 | 0.01411 | 0.17940 | 0.00649 | 0.21294 | 0.00119 |
|  | 2, 100 | 0.18912 | 0.00408 | 0.15988 | 0.01544 | 0.17353 | 0.00850 | 0.21700 | 0.00095 |
|  | 3, 150 | 0.18241 | 0.00563 | 0.15405 | 0.01968 | 0.16479 | 0.01252 | 0.21413 | 0.00111 |
|  | 4, 200 | 0.17873 | 0.00669 | 0.15050 | 0.02273 | 0.15935 | 0.01580 | 0.21265 | 0.00121 |
|  | 5, 250 | 0.17684 | 0.00731 | 0.14717 | 0.02594 | 0.15952 | 0.01568 | 0.21516 | 0.00105 |
|  | 6, 300 | 0.17425 | 0.00822 | 0.14533 | 0.02788 | 0.15929 | 0.01584 | 0.21344 | 0.00115 |
|  | 7, 350 | 0.17355 | 0.00849 | 0.14315 | 0.03035 | 0.16230 | 0.01393 | 0.21671 | 0.00096 |
|  | 8, 400 | 0.18416 | 0.00518 | 0.15420 | 0.01956 | 0.16884 | 0.01049 | 0.22583 | 0.00057 |
|  | 9, 450 | 0.19185 | 0.00356 | 0.16368 | 0.01313 | 0.17434 | 0.00819 | 0.22900 | 0.00048 |
|  | 10, 500 | 0.23517 | 0.00033 | 0.21582 | 0.00101 | 0.22330 | 0.00066 | 0.26950 | 0.00004 |
| **Variable** | **Measure** | **SRS total** | | **SRS communication** | | **SRS motivation** | | **SRS mannerisms** | |
| Age | Rho | 0.1755 | | 0.1475 | | 0.1606 | | 0.2137 | |
|  | *P*-value | 0.0079 | | 0.0260 | | 0.0152 | | 0.0012 | |
| Sex | Rho | 0.1402 | | 0.1065 | | 0.1322 | | 0.1880 | |
|  | *P*-value | 0.0344 | | 0.1088 | | 0.0462 | | 0.0044 | |
| Motion | Rho | 0.1644 | | 0.1361 | | 0.1446 | | 0.2028 | |
|  | *P*-value | 0.0129 | | 0.0400 | | 0.0291 | | 0.0021 | |

Supplemental Table 6. Assessing generalizability of the ADOS network in ABIDE using different network sizes and when partialling out different variables. Top rows: Testing different ADOS network sizes and determining prediction accuracy in ABIDE for SRS scores. The parameters tested and network sizes columns are the same as that used for Supplemental Table 5. Bottom rows: Testing prediction of SRS scales using the ADOS network and adjusting for different variables via partial correlation. All results: Rho = Spearman’s rho of the correlation between predicted and actual SRS scores. *P*-values (associated with the corresponding Spearman’s rho) for the alternative network sizes and when partialling out different variables are shown for context. Note that due to the post-hoc nature of this analysis, we are not interested in statistical significance of the new CPM pipelines per se. Rather, we are interested in the overall pattern of results and determining if results generally converge across the different pipelines, consistent with the multiverse approach used elsewhere in the paper^39^.. SRS, social responsiveness scale.

Testing different ADOS network sizes to predict SRS scores in HBN

|  |  |  |  | **SRS total scores** | |
| --- | --- | --- | --- | --- | --- |
| **Parameters tested** | **Positive network size** | **Negative network size** | **Combined network size** | **Rho** | ***P*-value** |
| 1, 50 | 5966 | 5812 | 11,778 | 0.0767 | 0.0521 |
| 2, 100 | 3119 | 3051 | 6,170 | 0.0757 | 0.0551 |
| 3, 150 | 2181 | 2176 | 4,357 | 0.0839 | 0.0335 |
| 4, 200 | 1670 | 1651 | 3,321 | 0.0862 | 0.0289 |
| 5, 250 | 1313 | 1261 | 2,574 | 0.0906 | 0.0217 |
| 6, 300 | 1001 | 1013 | 2,014 | 0.1002 | 0.0111 |
| 7, 350 | 773 | 763 | 1,536 | 0.0984 | 0.0126 |
| 8, 400 | 532 | 533 | 1,065 | 0.0923 | 0.0193 |
| 9, 450 | 283 | 277 | 560 | 0.0989 | 0.0121 |
| 10, 500 | 26 | 30 | 56 | 0.0423 | 0.2841 |

Supplemental Table 7. Testing different ADOS network sizes and determining prediction accuracy in HBN for SRS scores. Parameters tested and network size columns are as above. Rho = Spearman’s rho of the correlation between predicted SRS scores and actual SRS scores. *P*-values (associated with the corresponding Spearman’s rho) for the alternative network sizes are shown for context. Note that due to the post-hoc nature of this analysis, we are not interested in statistical significance of the new CPM pipelines per se. Rather, we are interested in the overall pattern of results and determining if results generally converge across the different pipelines, consistent with the multiverse approach used elsewhere in the paper^39^

Testing for convergence of task versus rest CPM findings in HBN: using the consensus model from HBN SRS prediction to predict ADOS scores in the Yale youth sample

|  | | | | **Results using combined network** | |
| --- | --- | --- | --- | --- | --- |
| **Parameters tested (folds, iterations)** | **Positive network size** | **Negative network size** | **Combined network size** | **Rho** | ***P*-value** |
| 1, 50 | 5,310 | 5,369 | 10,679 | 0.26 | 0.0413 |
| 2, 150 | 1,855 | 1,797 | 3,652 | 0.26 | 0.0382 |
| 3, 250 | 940 | 893 | 1,833 | 0.26 | 0.0422 |
| 4, 350 | 441 | 389 | 830 | 0.27 | 0.0361 |
| 5, 450 | 73 | 69 | 142 | 0.17 | 0.1832 |

Supplemental Table 8. Testing different DM consensus network sizes and determining the relationship between predicted SRS score and ADOS in the Yale youth sample. As above, we generated a variety of summary networks in the HBN sample (generated from the prediction of SRS) and assessed generalizability to predict ADOS in the Yale youth sample. To generate each summary network, we required that an edge appear in *n*/5 k-folds and *j*/500 CPM iterations (leftmost column). Specifically, the variables *n* and *j* were initially set at 1 and 50, respectively. The variables were altered in increments of 1 and 50, respectively. The rows of the table are arranged such that the top row is the most liberal network summarization approach (i.e., an edge had to appear in only 1/5 folds and 50/500 iterations); the bottom of the table is the most stringent case (i.e., an edge must appear in 5/5 folds and 450/500 iterations). Rho = Spearman’s rho of the correlation between predicted SRS scores and ADOS scores. *P*-values (associated with the corresponding Spearman’s rho) for the alternative network sizes are shown for context. Note that due to the post-hoc nature of this analysis, we are not interested in statistical significance of the new CPM pipelines per se. Rather, we are interested in the overall pattern of results and determining if results generally converge across the different pipelines, consistent with the multiverse approach used elsewhere in the paper^39^. Note models that included more edges (generated using a more liberal threshold) generalized, as did models generated using fewer edges (generated using more stringent thresholds). Also note that the only model that did not successfully generalize was the most stringent model comprising 142 total edges (~0.4% of the connectome), in line with results in Supplemental Tables 5 and 7 and noted in previous work^18^.

**Supplemental Discussion**

Neuroanatomy of the ADOS consensus model

In Supplemental Figure 1E, we present networks harboring predictive edges in the ADOS consensus model without correcting for network size. Similar to the data shown in the main text in Figure 6, both the positive-association and negative-association networks comprise a complex collection of edges, with representation within and between network pairs across the brain, with edges from across the brain. Also similar to the data shown in Figure 6, edges involving heteromodal association cortices contained the highest number of edges. In the positive network, connections linking the medial frontal to the motor network contained the most edges (107/1,001 edges). The next highest network pair (63/1,001 edges) comprised connections linking frontoparietal to motor nodes, while the third (48/1,001 edges) comprised connections between the medial frontal and visual association networks. In the negative network, connections linking the medial frontal and default mode networks contained the highest number of edges (75/1,013 edges). The next two highest network pairs comprised within-network medial frontal connections, and connections linking the medial frontal to the frontoparietal network (63/1,013 and 60/1,013 edges, respectively. That the heteromodal association networks again emerged as the top network pairs reinforces their importance in mediating the brain-behavior relationship in the ADOS consensus model.

Arousal, sample size, and case-control prediction in autism predictive modelling studies

Here we offer some additional thoughts related to arousal and sample size in the current work. It must be noted that the SSA task and resting-state conditions might have underperformed with respect to prediction performance as they came later in the study (i.e., following gradCPT) and participants were possibly fatigued. Nevertheless, previous work has shown that arousal does not appear to drive differences in predictions across different scanning conditions and that increased prediction performance seems to be due to cognitive differences driven by the task^35^. Future work could more fully investigate the role of arousal in brain-behavioral relationships in autism, while accounting for the challenging realities of scanning youth with neurodevelopmental conditions.

Much has been written about sample size in fMRI studies, and the need for both an adequate sample size and adequate data collection spanning modalities, allowing multi-modal brain-phenotype relationships to be studied. In this work, we attempted to balance a ‘deep’ approach, which involves collecting many scans from the same subject^40^, with a ‘broad’ approach, which involves collecting data from many subjects^34,41^. Hence, the data allowed us to compare the conditions across subjects, though this necessarily limited our sample size. Additionally, acquiring high-quality data from youth with autism is often time-intensive. In our experience, it is necessary to conduct a mock scan for at least an hour to adequately prepare a participant for the scanning environment^1^. The time commitment is on par with that reported by other groups in youth with neurodiverse conditions^27,42^.

In the main text, we discuss the notion of overlapping networks mediating attention and autistic phenotypes. We also consider the concept of the p-factor^43^. Directly in contrast to the idea that there is a single factor unifying broader pathology is the idea of distinct categories of functioning, typified by case-control classification studies^44-46^. Using the gradCPT data, we could successfully perform binary prediction of autism diagnosis. This is unsurprising, given that gradCPT resulted in the best dimensional prediction performance of ADOS scores, and autism diagnosis is related to higher ADOS scores (indeed, a high ADOS score is typically one component of helping to diagnose autism). In our view, successful case-control classification does not invalidate the use of dimensional approaches to predict the same phenotype. Considering that autism spans a spectrum of functioning, it is perhaps more biologically informed to use a dimensional approach at times, especially when attempting to make sense of participants who have a moderate degree of autistic traits. In other scenarios—for instance, trying to determine if a young participant has autism^45^, so they could be referred to the appropriate behavioral resources to improve adaptive functioning—a simple ‘yes’ or ‘no’ might be desired. While it is not currently feasible to scan infants to predict autism diagnoses in the future, encouraging progress has been reported^45^.
